# Priorities to Commitments: A binational adapted-NGT for refugee health prioritization

**DOI:** 10.64898/2026.10.26.26366541

**Authors:** Ana Cristina Sedas, Pere Kennedy, Mohammad Yasir Essar, Eric Norrie, Nesma El-Shazly, Myles Leslie, Sarah Clarke, Deyana Altahsh, Annalee Coakley, Gabriel E. Fabreau

## Abstract

**Background:** Priority-setting exercises shape research and policy agendas, yet their design is rarely reported, and the people most affected seldom participate. We present an adapted nominal group technique (NGT), embedded within an appreciative inquiry framework, where communities, professionals, and government set priorities together, voting separately on each priority’s importance and actionability.

**Methods:** The University of Calgary partnered with Immigration, Refugees and Citizenship Canada to host a two-day workshop (Calgary, July 2023) that convened clinicians, settlement providers, policymakers and academics from six Canadian provinces, seven US states and international organizations. It spanned three resettlement phases: pre-migration to early arrival, post-migration (up to two years), and long-term resettlement. A pre-workshop survey asked respondents to identify problems, what was working, and proposed solutions during each of these three phases. Solutions were grouped into preliminary solution lists. Participants expanded the lists, ranked solutions by weighted ballot, deliberated the leading priorities, and voted separately for the most important and most actionable priority in each phase. Survey responses and station notes were analysed qualitatively.

**Results:** Fifty-three of 68 participants completed the preworkshop survey. Participants generated 141 candidate solutions (73 from the survey and 68 at the workshop), which weighted ranking reduced to five priorities per phase. In two phases, the priority voted most important differed from the one voted most actionable: in Phase 1, a stakeholder taskforce was most important (49.0%) and linking pre-departure medical examinations to refugee clinics most actionable (64.7%); in Phase 2, normalizing refugee health care in primary care was most important (69.2%) and health navigation most actionable (47.9%). In Phase 3, surge readiness was most important (40.0%) and tied for most actionable (39.6%). Mental health dominated Phase 2 survey responses and funding recurred across all phases, yet neither was voted most important or most actionable in any phase.

**Conclusion:** An adapted NGT can build consensus across sectors that rarely deliberate together. Double-criterion voting separates importance from actionability, which a single ranking cannot. Pairing expert facilitation with research design offers a replicable model for whole-of-government, whole-of-society priority-setting; realizing its promise requires extending participation into process design and accountability for enacting jointly set priorities.

## Background

Priority-setting exercises guide how limited resources are directed toward research and health system improvement (1). Because evidence alone rarely determines action, they require transparent, rigorous processes to legitimize their findings (2,3). Historically, priority-setting has relied on expert panels deliberating separately, largely without the refugees and migrants whose care is being decided (4,5) and published exercises report their outputs rather than their process: who was included, how power imbalances were managed, and whether the approach can be reproduced (5). This matters because who is convened, and how they deliberate, shapes what is prioritized.

Nominal group technique (NGT) is a structured consensus method that moves from silent idea generation to ranking, equalizing participation and producing a ranked list quickly. NGT has been widely used in healthcare (6,7), including by our team with a refugee patient advisory committee across three phases of resettlement (8). Struggling to represent diverse stakeholders (9,10), ranked lists that do not capture the nuance in participants’ ideas and ranking on a single criterion that conflates importance with feasibility, which often diverge when rated separately (11), are known limitations of NGT (12). Appreciative inquiry, a strengths-based approach to change, begins from what already works, an existing data-sharing platform, a health navigator role, a partnership that functions, and asks how it can be built upon rather than starting from what is absent (13). Embedding NGT within this framework shifts deliberation away from simply naming problems and toward designing solutions that can be implemented and scaled (14,15).

Refugee health especially needs such an approach, because responsibility for refugees’ care is divided across immigration, health and settlement sectors and across levels of government (16). With over 117.8 million people were forcibly displaced worldwide in 2025 (17). Canada and the United States, have received record arrivals between 2021 and 2023. For Canada, historic numbers of refugees, warranted a rapid response for Afghan refugees and Ukrainian evacuees, receiving more than 144,000 asylum claims in 2023. Receiving health systems were unable to keep up the pace (18,19). Yet practical, replicable mechanisms for cross-sector priority-setting, consistent with the WHO’s ‘whole-of-government, whole-of-society’ approach, remain scarce (20).

Priority-setting in refugee and migrant health runs on two tracks that rarely meet. One sets research priorities with strong clinical and community input (8) but limited translation into health system or policy change. The other sets priorities in forums convened by governments, academia and international agencies, such as the World Health Organization’s global research agenda on health and migration (21) and its regional adaptations (20,22,23). Across such exercises, translation into implementation plans is rarely addressed and evidence of change in clinical practice is rarely reported (24). These forums debate health and immigration policy at the system level but seldom include the clinicians and settlement workers who deliver care, so the priorities they produce rarely reflect the day-to-day of receiving or delivering it. Building on the participatory nature of NGT, we convened those who make decisions, those who implement them, and those affected by them at a two-day workshop in Calgary, Canada. To our knowledge, this combination has not previously been reported in North American refugee health policy priority-setting. We aimed to: (1) identify consensus-based priorities that are both important and actionable for refugee and migrant health policy in North America across three phases of resettlement; (2) describe the application of an adapted NGT within an appreciative inquiry framework as a structured approach to multistakeholder priority-setting; and (3) trace how priorities moved from survey through ranking, deliberation and the final vote, to identify what each NGT adaptation contributed to the final set of priorities.

## Methods

### Study design, theoretical framework and phases of resettlement

We conducted a multimodal qualitative policy consensus study using an adapted NGT embedded within an appreciative inquiry framework and grounded in pragmatist and participatory epistemological stances (25).The study had two parts: a pre-workshop online survey (Part 1) and a two-day workshop in Calgary, Canada, on July 19-20, 2023 (Part 2; Figure 1). We retained the logic of the five classic NGT steps (problem presentation, silent idea generation, round-robin sharing, group discussion and clarification, and ranking) but adapted their delivery: Steps 1 and 2 were conducted through the survey; at the workshop, silent generation was repeated to expand the survey-generated list, followed by discussion and clarification (Step 3), weighted ranking (Step 4), progressive consensus building (Step 5a), and a final vote on importance and actionability (Step 5b). Steps 3 to 5a were repeated for each resettlement phase; Step 5b covered all three. Because quantitative outputs (survey ratings, rankings, and votes) were summarized descriptively and no data were analyzed by stakeholder group or jurisdiction, we describe the design as multimodal qualitative rather than mixed methods.

**Figure 1.**
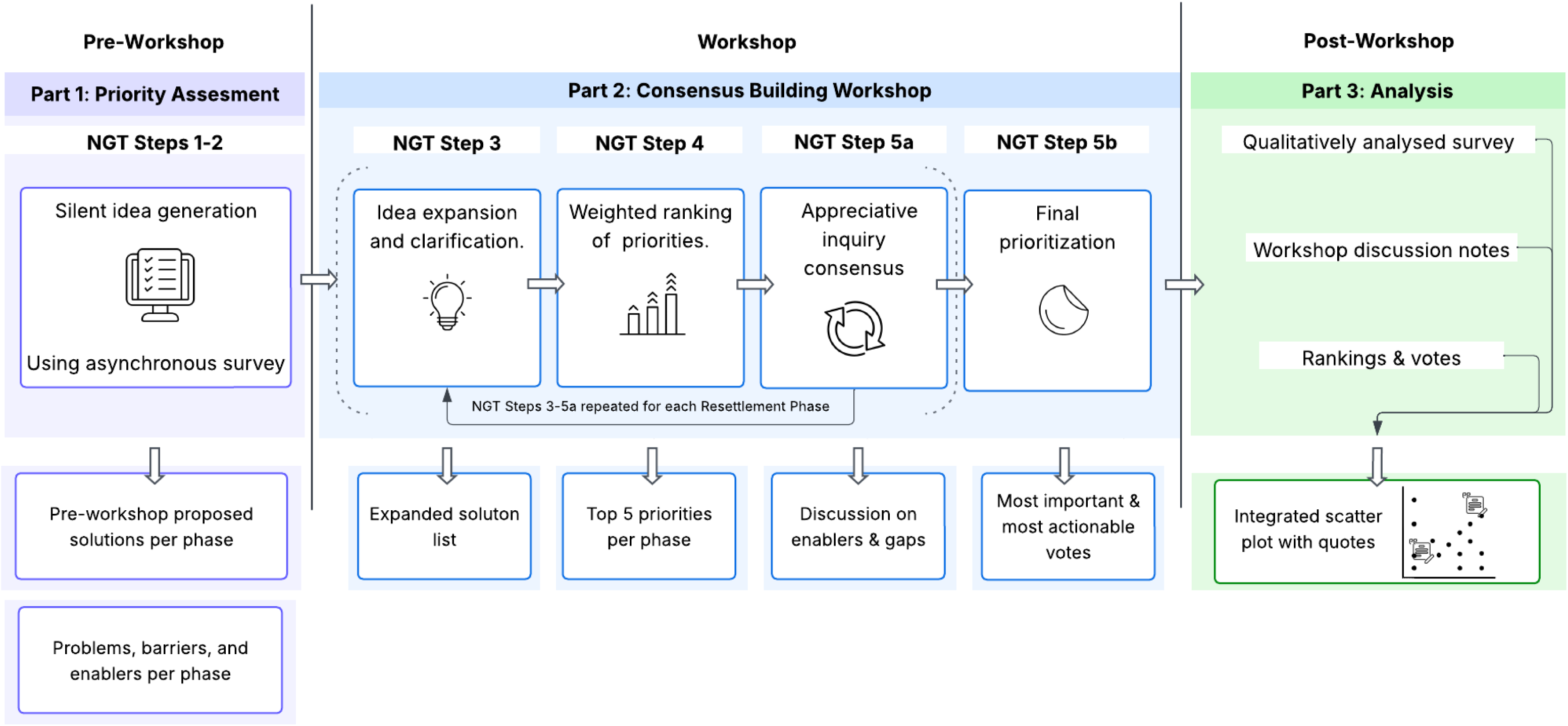
Visual depiction of the study’s procedure. NGT = Nominal Group Technique.

We organized the study across three resettlement phases (hereafter Phases 1, 2, and 3), adapted from our previous NGT study with a refugee patient advisory committee (8) and set a priori as operational periods of healthcare access. Phase 1, pre-migration to initial post-arrival, spans three months before departure to three months after arrival, aligning with United States resettlement policy that expects economic independence 90 days after arrival (8). Phase 2, early post-arrival, spans three months to two years after arrival, the period most specialized Canadian refugee clinics care for newly arrived refugees for one to two years post resettlement (8,26). Phase 3, long-term resettlement begins two years after arrival and was examined through the lens of refugee sensitive health system capacity, surge preparedness, and sustainability for long-term health.

The University of Calgary Conjoint Health Research Ethics Board approved the study (REB 18-2074). Survey completion after a waiver of consent was taken as implied consent; workshop participants provided verbal consent. We followed the Reporting Guideline for Priority Setting of Health Research (27).

### Study setting and participant selection

The workshop was co-hosted by the University of Calgary’s Migration Humanitarian Health Collective (formerly Refugee Health YYC), O’Brien Institute for Public Health, and School of Public Policy, in partnership with Immigration, Refugees and Citizenship Canada (IRCC) and the Society of Refugee Healthcare Providers (SRHP), as a pre-conference to the 2023 North American Refugee Health Conference. We used purposive and network sampling to invite refugee community leaders, clinicians, academics, public health officials, policymakers, and resettlement service providers from government, civil society, and international organizations through established networks and conference registrants; invitations were sent by email between April and June 2023.

### Part 1: Baseline priority assessment (NGT Steps 1-2)

The survey (Qualtrics, Provo, UT) (28) was sent to all invitees with their workshop invitation and comprised sociodemographic items and questions on each resettlement phase. We defined “refugees” to include forcibly displaced populations, including refugees, asylum claimants, and Ukrainian evacuees. Consistent with appreciative inquiry, each phase’s section opened with one or two closed items asking respondents to rate access or capacity on a 0–10 scale, followed by open-ended questions asking them to describe problems and barriers, identify what works well, and suggest how successes could be scaled and what solutions could improve care (Additional File 2). The survey produced two kinds of data: numerical ratings, which we summarized descriptively, and unstructured written responses forming the qualitative data set. Responses were matched by name to the workshop attendance list, assigned a unique study ID, and de-identified; repeat submissions were merged. From the open-ended responses to the solutions questions, the research team grouped similar proposed solutions and merged duplicates to form a preliminary solution list for each resettlement phase, which was presented at the workshop.

### Part 2: Consensus-building workshop (NGT Steps 3-5)

An experienced NGT facilitator led the workshop, co-designed with refugee and migrant health experts and opened by refugee community leaders sharing their experiences of displacement and resettlement. We addressed each phase in turn through Steps 3 to 5a over two days, each accompanied by case presentations of current practices across jurisdictions, followed by final prioritization (Step 5b) at the end of Day 2.

### Step 3: Priority expansion and clarification

In plenary, participants first reviewed the preliminary survey-generated solutions for each phase, then generated additional solutions silently and individually on paper at their tables, repeating the NGT silent-generation step. A research team member added these to the list in real time with full group visibility, and participants asked clarifying questions about any solution on the expanded list before ranking.

### Step 4: Weighted ranking

Participants independently ranked their top five solutions for each phase by confidential paper ballot, first to fifth choices receiving five to one points (maximum 15 points per participant per phase), and the research team tallied ballots in Microsoft Excel. After tallying, the facilitator proposed combining solutions generated separately that described the same action, and participants agreed in plenary which to merge; a merged priority took the rank of its highest-scoring component, and we report each component’s points. Where two solutions tied at fifth, participants decided as a group which to retain. The five highest-scoring priorities per phase (15 overall) went forward to Steps 5a and 5b; all 141 solutions are listed with their points in Additional files 1a–1c.

### Step 5a: Progressive consensus building

Each leading priority was assigned to a station. Small groups rotated at 15-minute intervals through three stations, where flip charts structured discussion around three appreciative inquiry questions: (1) what is working well and can be leveraged; (2) what is not working well and how it can be improved; and (3) how integration and coordination can be enhanced. Groups wrote their responses on the flip chart and marked earlier groups’ entries with a checkmark where they agreed or an X where they disputed; a research team member later transcribed these verbatim, forming the only verbatim record of participants’ reasoning. Nominated delegates summarized each station in plenary, followed by debate and member checking.

### Step 5b: Final prioritization

The research team presented the 15 leading priorities in a final plenary, one flip chart per phase, and participants shared observations across phases. Final prioritization used dot voting, a common voting method in NGT. Each participant received three yellow and three blue dots and was asked to place one yellow dot on the priority they considered most important and one blue dot on the priority they considered most actionable in each phase; a participant could select the same priority for both. Limiting each participant to one dot of each color per phase prevented a few participants from concentrating dots on a single option. Because not all participants distributed their dots as instructed, dot totals varied by phase and category.

### Quantitative and NGT analysis

Closed-ended survey items were summarized as counts and percentages, or mean and standard deviation for age, with “prefer not to disclose” treated as missing and not imputed. The 0-10 ratings are summarized as medians (IQR) and the proportion rating 5 or lower.

For Step 4, solutions were ordered by total weighted points; because some ballots ranked fewer than five solutions and a few appeared to have repeated a rank, we took the number of ballots per phase as the largest number of votes cast at any single rank. The final vote (Step 5b) was the principal output: for each priority, we report its share of yellow (most important) and blue (most actionable) dots and compare importance with actionability, and both with the weighted ranking (Table 2; Figure 2).

**Figure 2.**
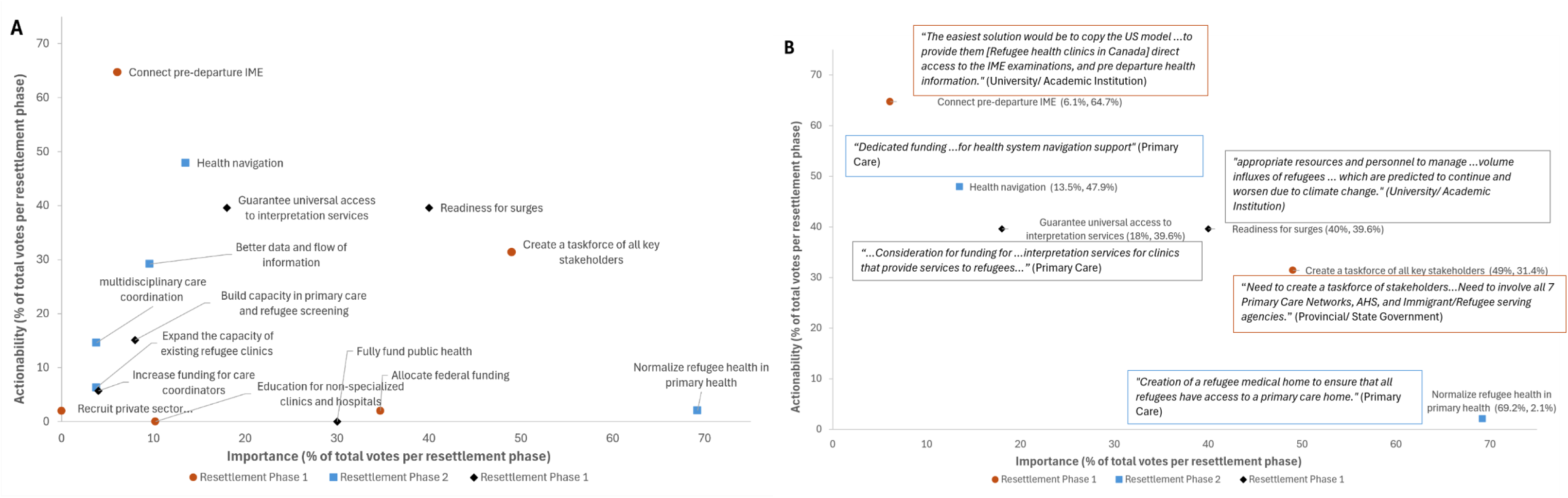
Importance and actionability of the 15 leading priorities in the final vote, by resettlement phase. (A) Share of most-important votes (x-axis) and most-actionable votes (y-axis) for each Priority. Denominators were 49 and 51 dots for Phase 1 (pre-migration to initial arrival; orange circles), 52 and 48 for Phase 2 (post-migration; blue squares), and 50 and 53 for Phase 3 (long-term resettlement; black diamonds). (B) The most important and most actionable priorities in each phase, with illustrative quotes from survey respondents. Priority titles are shortened in both panels.

### Qualitative analysis

After the workshop, the research team developed a codebook to analyse the open-ended survey responses generated during NGT Steps 1-2 (Additional file 3). Stance (problem, barrier, enabler, proposed solution, or context) were assigned deductively from a priori developed definitions themes were developed inductively from the data. If certain pieces of text matched a different resettlement phase than originally described, this was also identified. Two coders double coded all responses, working independently and resolving disagreements through discussion until consensus. The qualitative analysis was not part of the workshop process and did not inform the preliminary solution list, the ranking, or the final vote; rather, it allowed the research team to trace the problems, barriers, and enablers respondents described to the priorities participants later ranked and voted on.

Workshop responses from Step 5a were not coded. Because they sit between the weighted ranking and the final vote, we used them to illustrate the reasoning that led participants to their final choices, rather than as material for coding. Where these responses appear in the paper, they are attributed as “Workshop participant”

### Data Integration

We integrated three data streams in Figure 2, which pairs a scatter plot of importance and actionability votes with illustrative quotes: the survey (ratings and coded open-ended responses), which showed what respondents raised in each phase; the consensus-station responses, which showed how participants reasoned about each priority; and the weighted rankings and final votes, which traced how priorities moved through the process. Where streams diverged, we did not reconcile them but used the divergence to identify priorities formed by consolidation rather than raised in the survey, and priorities voted on in ways the ranking did not predict.

### Researcher reflexivity

The senior authors and several co-authors (GF, EN, NES, ML, AC) designed, hosted and/or attended the workshop. The lead analyst and the second and third authors (ACS, PK, MYE), who developed the codebook, coded the data and conceptualized this manuscript, did not. Analyzing the data without having attended reduced the risk that recollection of the room shaped the analysis, but it also meant the analysts had access only to what was written (the verbatim station responses and the ballot tallies), not to the exchanges that produced them or the plenary debates, which were not recorded. Attending co-authors reviewed Table 2, Figure 2 and the account of consolidation against their recollection, with disagreements resolved by returning to the source data.

## Results

### Participant characteristics

Sixty-eight participants attended the workshop: 44 (65%) attended both days, 15 (22%) Day 1 only, and 9 (13%) Day 2 only, giving 59 participants on Day 1 and 53 on Day 2 (Table 1). Fifty-three people responded to the baseline survey, including 45 workshop participants (66%) and eight invitees who did not attend. Respondents’ mean age was 45 years (SD 11), and 31 (58%) identified as female Most worked in Canada (30; 57%) or the United States (15; 28%); eight (15%) were based elsewhere or did not report a country. Respondents worked predominantly in primary care (17; 32%), universities or academic institutions (12; 23%), government agencies (8; 15%), and public health (5; 9%), with settlement agencies and other organizations (6; 11%) also represented; five (9%) did not report an organization type. Thirty-one respondents (58%) reported interacting with refugees daily or weekly and 18 (34%) monthly, rarely, or never; four (8%) did not report (Table 1).

**Table 1.**
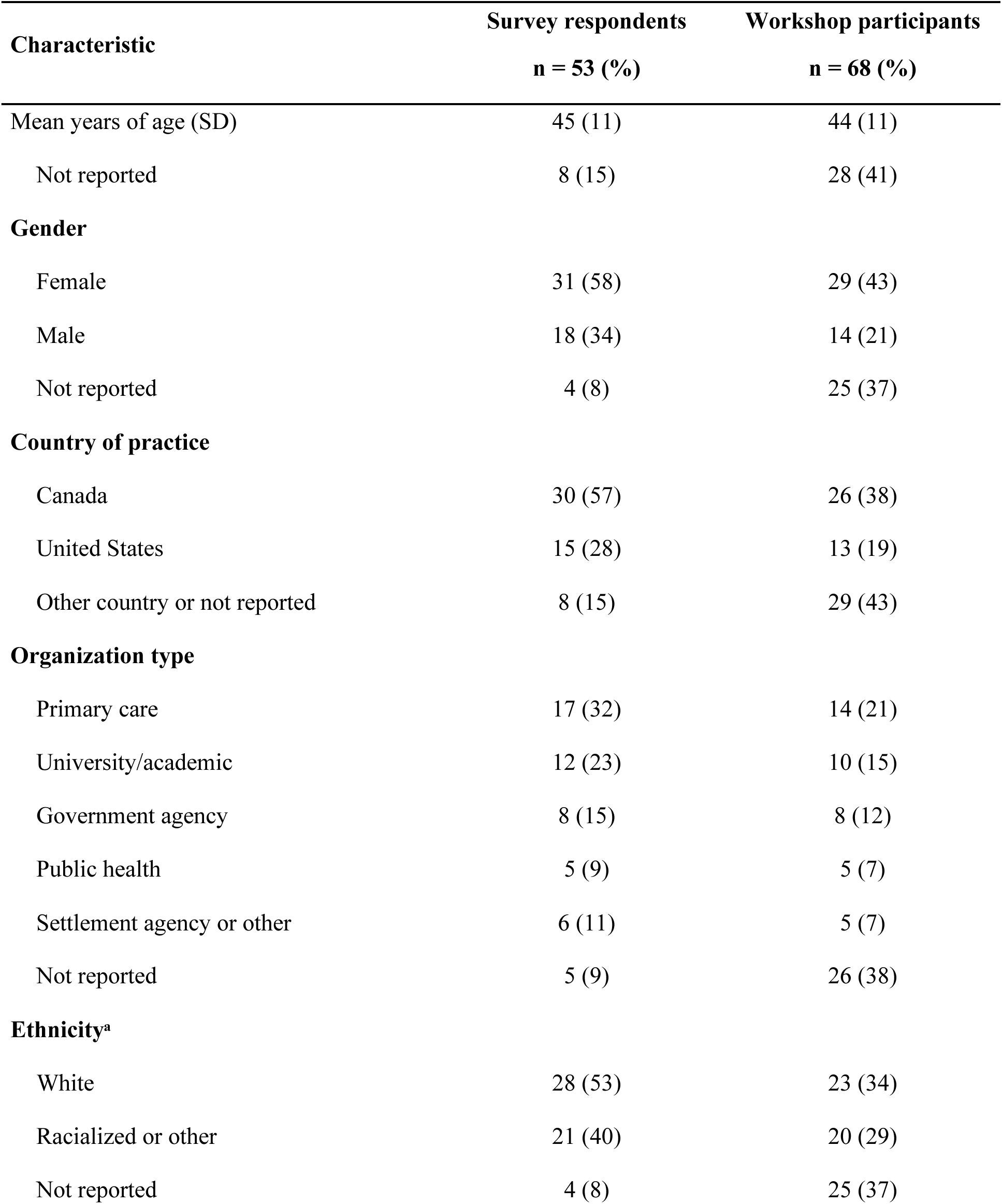
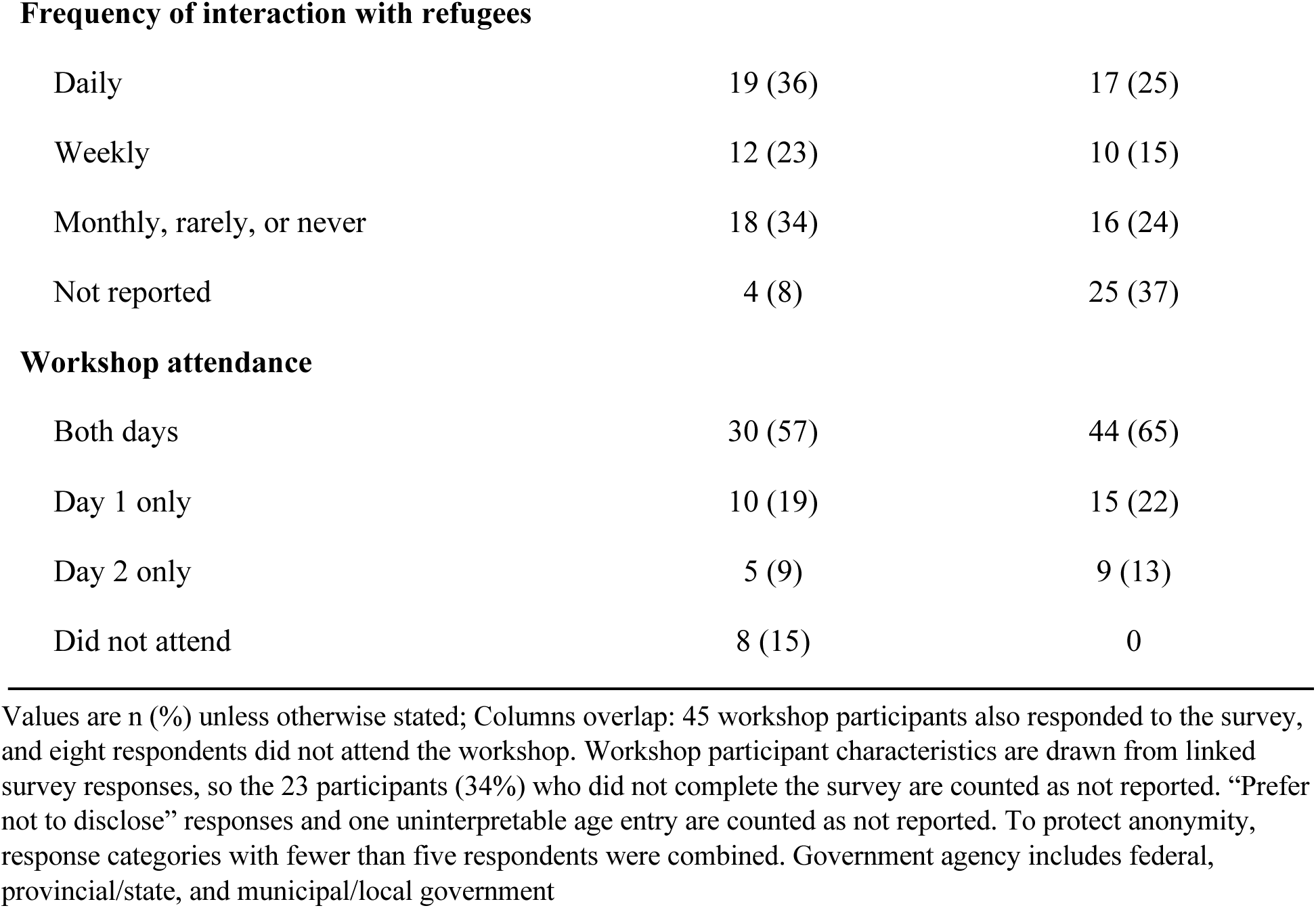
Characteristics of survey respondents and workshop participants.

**Table 2.**
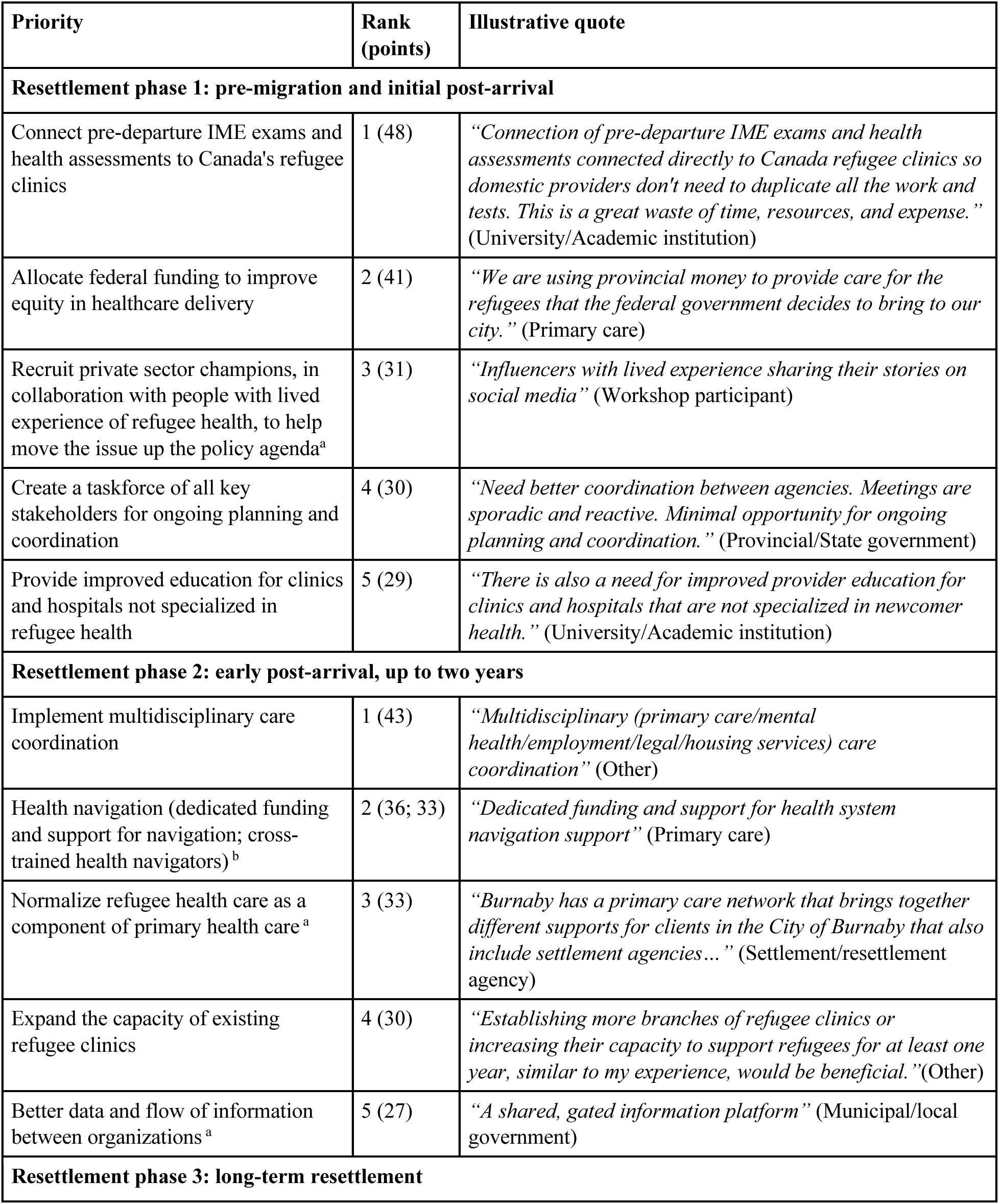
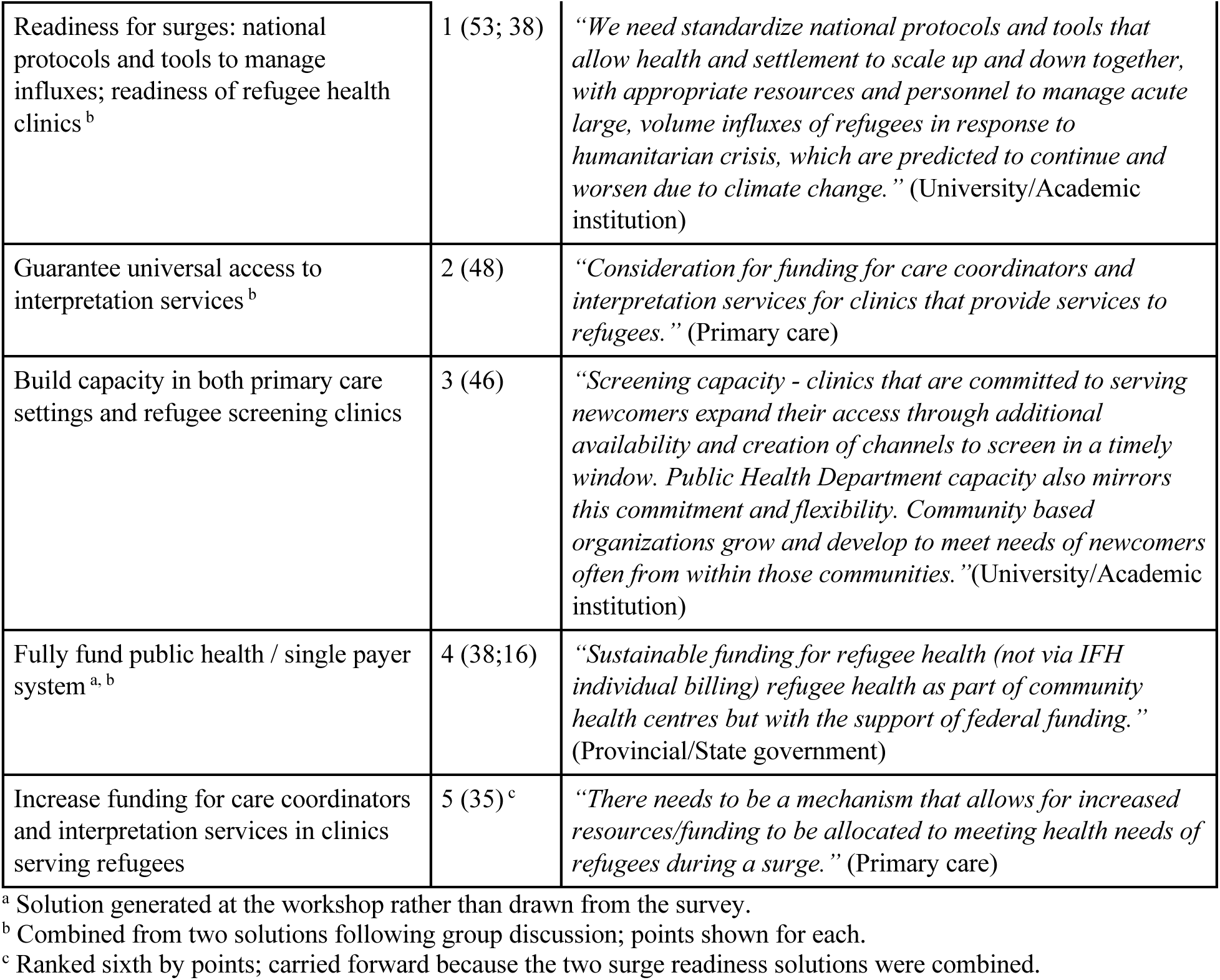
Five leading priorities per resettlement phase after weighted ranking (Step 4), with rank, total points and an illustrative quote.

## Part 1: Baseline Priority Assessment

### NGT Steps 1 and 2. Silent Idea Generation

#### Resettlement Phase 1 – Pre-Migration and Initial post-arrival

Survey respondents rated domestic access to pre-departure health assessment information as poor (n = 43; median 2 of 10 [IQR 1-6]), with 31 (72%) rating access 5 or lower.

Pre-departure medical information, including immigration medical examinations (IMEs) and health assessments, rarely reached post-arrival health clinics. Respondents attributed this to the absence of formal channels between pre-departure and resettlement actors and described the gap as more pronounced amongst asylum seekers than government-supported refugees. Respondents in the United States, however, pointed to the repurposing the CDC’s (Centre for Disease Control) Electronic Disease Notification System in partnership with the IOM (International Organization for Migration) as an enabler; giving providers access to pre-departure exams and complete records before arrival.

*“It appears that whatever information is available is accessible for the settlement agency, but does not necessarily get transferred to refugee health clinics.”* (Primary care)

Respondents further reported a lack of formal coordination between federal immigration agencies and state/provincial clinics; describing the system as fragmented. Respondents identified limited funding as a central problem in providing specialized refugee care, particularly during the acute integration phase. Limited funding left specialized clinics and settlement agencies with staffing constraints; outside specialized services, care relied on staff not sufficiently trained in refugee health.

#### Resettlement Phase 2 – Early post-arrival ( up to two years)

Survey respondents rated the accessibility of mental health services for newly arrived refugees as low (n = 39; median 3 [IQR 2.0-5.5]), with 29 (74%) rating it 5 or lower.

Respondents described problems similar to Phase 1: coordination between sectors remained a challenge, with little delineation of roles and responsibilities between health and resettlement agencies; clinics and settlement agencies were often underfunded and lacked the bandwidth to coordinate efficiently.

Access to general medical services was difficult, mainly because of challenges navigating the system. Some respondents also described racism in care, with providers more willing to support refugees from some countries than others.

*“Casual racism is a concern as well (e.g. many providers were more willing to offer support for Ukranians and less so for Afghans).”* (Provincial/state government)

Unique to this phase, respondents often described limited access to mental health services for refugees and newcomers. Mental health and counselling capacity was limited and, where available, rarely culturally sensitive.

*“In our jurisdiction we have worked closely with a private clinic which provides psychotherapy…and counseling which bills IFHP [Interim Federal Health Program] and is comfortable using interpretation. This is new…before this we did not have any private psychotherapy options.”* (Primary care)

#### Resettlement Phase 3 – Long-term resettlement

Survey respondents rated the capacity of existing health and settlement services as insufficient relative to both current arrivals (n = 34; median 4 [IQR 2-5]), with 26 (76%) rating capacity 5 or lower, and projected arrivals (n = 33; median 4 [IQR 2-5]), with 25 (76%) rating capacity 5 or lower.

Respondents’ problems and barriers centered on the capacity to meet current needs and on continuing surges of arrivals. Respondents highlighted limited human and financial capacity to meet needs, compounded by staff burnout, task-shifting beyond their roles, and high turnover. Wait-times lengthened as a result, and were more pronounced among privately sponsored refugees, claimants and those arriving through special migration pathways.

*“I think all areas have been struggling to respond as resources have come behind the need….”* (Settlement/Resettlement Agency)

Budgets did not keep up with arrivals, constraining staffing. Where resources existed, poor coordination produced short-term solutions to long-term problems.

*“We are currently only funded for 1NP [Nurse Practitioner] and 1RN [Registered Nurse] for refugee health from the province. All other resources are ‘borrowed’ from the community health centre…we are forced to turn down supporting many others who are in need…”* (Primary care)

Respondents highlighted the need to create local systems led by local governments and in close partnership with clinics and local communities. Respondents explored standardizing protocols and tools that not only address immediate surges (preparedness and response plans) but support homogenization of good practices across the entire health system. See Additional files 5a-5C for all problems and barriers described by survey respondents.

### Part 2 – Consensus-building workshop

#### NGT Steps 3 and 4. Priority expansion and weighted ranking

Participants considered 141 candidate solutions: 60 in Phase 1 (35 from the survey and 25 added at the workshop), 43 in Phase 2 (18 and 25), and 38 in Phase 3 (20 and 18) (Additional files 1a-1c). Participants submitted 43, 46, and 48 ranking ballots in Phases 1 to 3, for a maximum possible 215, 230, and 240 points per solution. After tallying, the two navigation solutions (Phase 2) and the two surge solutions (Phase 3) were combined, and single-payer healthcare was combined with fully funding public health (Phase 3). Ties at the cutoff were resolved by group discussion: in Phase 1, improving provider understanding of trauma and cultural contexts (29 points) was consolidated with the education priority; in Phase 2, a mechanism to scale up health supports during surges (27 points) was not carried forward. Table 2 shows the five leading priorities per phase with their points, origin, and an illustrative quote.

#### NGT Step 5a: Progressive consensus building

At the consensus stations, participants examined each leading priority against what was already working, what was not, and what it would take to improve it, before voting on importance and actionability (Additional files 4a-4c).

In resettlement phase 1, participants identified the top-ranked priority, connecting pre-departure IMEs to refugee clinics, as a technical gap. Participants discussed the CDC’s Electronic Disease Notification system as “a replicable model” and listed concrete next steps including creating a unique identifier for arrivals. The taskforce, tied third, had both a working model (existing councils, convenors, an IRCC health liaison) and an actor present who could act on it (IRCC had co-hosted the workshop). Federal funding for equity, ranked second, had no working model to point to; its station notes asked for sustainable funding “s*pecific for health and migration rather than compete for other programs*” a demand on government rather than a task for anyone in the room. *“Funding should be specific for health and migration rather than compete for other programs.”* (Workshop Deliberation Participant)

This pattern is repeated in resettlement phase 2. Participants discussed specific existing models for health navigation; ranked second. The Minnesota clinic has existing navigators and counselling groups however, its gap was training and role clarity. Normalizing refugee health care within primary care, ranked third, yet its station entries were tools, “*websites and apps such as the Refugee Care App*”, rather than a route to primary care taking on refugee health as standard.

*“Use websites and apps such as the Refugee Care App (Canada) and CareRef (US) to provide resources for general practitioners”* (Workshop Deliberation Participant)

In phase 3, participants framed national protocols for surge readiness, the top-ranked priority, as both necessary and within the remit of the agencies represented. For interpretation, ranked second, participants pointed to an existing mechanism that could be extended, using Interim Federal Health Program billing to hire more providers, and it matched surge readiness on actionability. Across the phases, participants described a siloed, reactive system in which funding was disconnected from service provision.

*“Passion and interest in dedicated refugee clinic — good: work gets done; bad: burnout/unpaid work”* (Workshop Deliberation Participant)

#### NGT Step 5b: Final prioritization

Participants placed 49 yellow and 51 blue dots across the five leading priorities for Resettlement Phase 1, 52 yellow and 48 blue for Resettlement Phase 2, and 50 yellow and 53 blue for Resettlement Phase 3. In Phases 1 and 2, the priority ranked first was not the one voted most important, and in Phase 2 the priority voted most important was the least actionable.

Figure 2 plots each priority’s share of importance votes against its share of actionability votes; priorities toward the upper right were judged both important and actionable.

Connecting pre-departure IME (Immigration Medical Exams) and health assessments to post-arrival refugee clinics in Canada was voted as the most actionable priority in Resettlement Phase 1 (33/51, 64.7%) (Figure 2B). Multiple respondents discussed the importance of having patient information arrive in a timely manner to appropriately address the needs of new arrivals. During the workshop participants were made aware of current models of sharing patient information across clinics and agencies, such as the existing EDN (Electronic Disease Notification) system. Workshop attendants discussed the possibility to implement this US based model in Canada as well (Figure 2B).

Interestingly, despite numerous survey respondents discussing the need to improve information sharing, this priority was voted as the least important (Figure 2B).

Establishing health navigators was voted the most actionable priority in Phase 2 (23/48, 47.9%), and in Phase 3 guaranteeing universal access to interpretation services tied with surge readiness as most actionable (21/53, 39.6% each) (Figure 2B). Surge readiness was also voted most important in Phase 3 (20/50, 40.0%), the only priority to lead on both criteria, and it ranked first in the weighted ranking as well. In breakout discussions, participants framed national surge protocols as both necessary and within the remit of the agencies represented in the room, which may explain why importance and actionability aligned. Surge capacity had also been identified as a priority by IRCC partners during planning for the workshop and survey. Health navigation and interpretation, by contrast, both aim to make clinical and settlement services more accessible and easier to navigate for patients. Multiple survey respondents highlighted an urgent need for funding to establish such services in clinics and settlement agencies, yet participants voted both less important than the leading priority in their phase (navigation 7/52, 13.5%; interpretation 9/50, 18.0%) (Figure 2B).

All priorities voted as important (Creating a taskforce for ongoing planning and coordination (24/49, 49.0%), Normalizing refugee health in primary health (36/52, 69.2%), and Building surge readiness (20/50, 40.0%) (Figure 2B)); were discussed numerous times by various participants throughout the survey and workshop discussions. Additionally, all three require more structural changes to the North American Health system, which although important may require time and reform for actionability.

Three leading priorities (health navigation, surge readiness, and public funding) were formed by combining related solutions after ranking, and five were generated at the workshop rather than the survey (Table 2). Mental health, the problem most often described in Phase 2 survey responses, was not among the leading priorities in any phase, and funding priorities, although leading in Phases 1 and 3, were not voted most important or most actionable in either. Overall, participants judged discrete operational measures (IME-to-clinic data linkage, patient navigation, guaranteed interpretation) as most actionable, and structural reforms (a stakeholder taskforce, normalizing refugee care within primary care) as most important but less immediately actionable. Surge readiness bridged the two, voted both most important and jointly most actionable in Phase 3.

## Discussion

This study yields three primary insights. First, an adapted NGT made it possible to conduct rigorous, democratic priority-setting across national and subnational jurisdictions: over two days, policymakers, health professionals, settlement providers, and refugee community leaders deliberated as a single group within a structure designed to flatten power differentials. To our knowledge, this is the first time these constituencies have been convened together in North America for structured priority-setting across phases of resettlement, extending an approach previously applied with refugees themselves (8). Second, the priorities participants deemed most important were often not those they deemed most actionable. Third, priorities shifted across phases, with some, such as funding, cutting across all three. Survey respondents also described three barriers that persisted across all phases: limited data sharing between stakeholders, weak coordination and governance across health and migration systems, and limited, inconsistent funding.

Where priority-setting exercises rank on a single criterion, the priorities most likely to be acted on may be invisible: priorities are not judged on whether they can be implemented, and remain aspirational, contingent on funding and political will that may never arrive (29–31). When NGT exercises do consider actionability, it is often combined with importance, obscuring the distinction participants were able to draw in our study (30). Separating the criteria shows where work can begin and where advocacy is needed. Because the ranking and the final vote used different criteria, however, these shifts may reflect the criteria as well as the deliberation itself.

Tracing priorities from the survey through ranking, deliberation and the final vote shows that each adaptation contributed. First, the design operationalized the whole-of-government, whole-of-society approach that the UCL-Lancet Commission and the WHO Global Action Plan call for without specifying how to achieve it (32,33), bringing government, settlement providers, health professionals, and community leaders from Canada and the United States into one process. Second, conducting silent idea generation asynchronously through a pre-workshop survey drew in geographically dispersed perspectives, gave respondents time to reflect on each phase, and captured ideas without group influence; eight respondents who did not attend the workshop also contributed survey responses. Third, staging deliberation between ranking and voting, and asking participants to judge importance and actionability separately, changed which priorities rose to the top. Dot voting is common in NGT (12,34) and has been combined with impact-feasibility assessment in health priority-setting (35); voting on importance and actionability as separate criteria extends that approach. In Phase 1, the top-ranked priority, linking pre-departure medical examinations to refugee clinics, was deemed by far the most actionable (64.7%) but rarely the most important (6.1%), while a stakeholder taskforce, ranked fourth, was deemed most important (49.0%); in Phase 2, normalizing refugee health care in primary care, ranked third, was deemed most important by more than two-thirds of participants (69.2%). Funding followed the same pattern: federal funding in Phase 1 and fully funding public health in Phase 3 were often chosen as most important but almost never as most actionable. Participants appeared to distinguish operational fixes that existing actors could implement from structural reforms requiring political will and sustained funding.

Data sharing illustrates how these mechanisms worked together. Respondents described a lack of data sharing between stakeholders that affected both providers’ access to medical records and the quality-of-care patients received. Respondents practicing in Canada rated access to pre-departure health information far lower than those in the United States (median 2 vs 6 of 10), where the partnership between the CDC and the IOM, through the Electronic Disease Notification system, enables swift data transfer and early notification of arrivals (36). Participants carried this contrast through the workshop, considering how to replicate working models, and ultimately voted linking pre-departure examinations to refugee clinics as the most actionable priority of Phase 1.

The need for coordination and stronger health and migration governance emerged across all participant groups, in both the survey and the workshop. Accounts varied: some participants reported no formal coordination channels for routine arrivals, while others in the same country, and sometimes the same province, described functioning ones. Some arrivals were referred with complete documentation, but asylum seekers, people in detention, and privately sponsored refugees often were not. Consistent with previous literature, functioning coordination systems may reach only a subset of arrivals, leaving care teams unable to plan and shifting the burden of fragmented systems onto patients (37,38). Participants’ proposed remedy, a taskforce of all key stakeholders for ongoing planning and coordination, drew 49.0% of importance votes and 31.4% of actionability votes in Phase 1, one of only two priorities, with surge readiness, to receive substantial support on both criteria.

Funding emerged as a constant problem and, across every phase, as a precondition for many proposed solutions. Respondents described budget constraints preventing the retention of migrant-sensitive staff, clinics unable to employ health navigators or guarantee interpretation, and a one-year limit on Interim Federal Health Program coverage leaving people without mental health care. These concerns echo earlier findings that changes to the program generated confusion and shifted responsibility for refugee health to provinces (19,39) or, as one participant put it, provincial money paying for the care of refugees the federal government resettles. Because funding was attached to so many candidate solutions, it was repeatedly absorbed into broader priorities as the NGT progressed, suggesting that rank-based methods may underrepresent cross-cutting constraints; our appreciative inquiry questions helped make it visible.

Our findings extend evidence from our companion study, in which a diverse refugee patient advisory committee used NGT to prioritize health system navigation, and primary care capacity (8). The vertical alignment between priorities identified independently by resettled refugees at the community level and by multisectoral stakeholders at the policy level, particularly navigation, interpretation, and system capacity, strengthens the legitimacy and convergent validity of both agendas. Newcomer communities in our related work have likewise identified language access, navigation support, and trusted community intermediaries as preconditions for equitable care (8,40). Our priorities also overlap substantially with the WHO Global Research Agenda on Health, Migration and Displacement, including its themes of universal health coverage, emergency preparedness, and social determinants of health (21).

## Limitations

Our study has limitations that may inform future applications of this approach. First, the survey’s design may have shaped responses. The phases were set a priori, bundling pre-departure, a distinct stage, with early arrival. The survey’s focus also reflected priorities identified in consultation with the IRCC, including access to pre-departure health data, mental health services, and surge capacity, which informed the survey rating item and may have primed responses in each phase. However, this was partly addressed through solutions expansion. The survey’s framing may nonetheless have narrowed the range of issues respondents raised, and is relevant to interpreting why mental health, prominent in survey responses, did not reach the final vote. In addition, the survey asked about ethnicity but not migration background, so the participation of people with lived experience of migration is likely undercounted. Future iterations should treat pre-departure as its own phase, vary question order, and ask about migration background directly. Second, workshop discussions were not recorded and were captured only through notes and flip charts, limiting the detail available on why participants voted as they did. Voting was independent but not blind, and members of the organizing team were present, so social desirability and conformity may have influenced votes. Audio-recording deliberations and using private voting, such as paper ballots or anonymous digital polling, would address both limitations. Third, consolidating related solutions as the NGT progressed reduced the visibility of constraints that cut across several of them. In Phase 2, one solution tied at the cutoff was not carried forward to the final vote. Recording consolidation and tie-break decisions as they happen and confirming them with participants would make this step more transparent. Lastly, carrying five priorities per phase was a practical choice to fit two days; it does not imply that lower-ranked solutions lacked value, and several received substantial points.

## Conclusion

This study adapted the NGT for multistakeholder policy priority-setting by asking participants to judge importance and actionability separately, moving idea generation online, and embedding the process within an appreciative inquiry framework. Policymakers, providers, settlement workers, and refugee community leaders deliberated in one room and examined what was working before proposing what should change, showing that democratic, all-of-government, all-of-society priority-setting exercise is possible. The resulting priorities differed across phases of resettlement, and what participants deemed most important was often not what they deemed most actionable. Health systems serving refugees will need to respond accordingly, matching solutions to the phase of resettlement and beginning with actionable steps while building the conditions for the reforms participants valued most. Priorities are realized when decision-makers commit to them and agree on who will do what, by when, and with what resources. Future applications could build that step into the process itself, so that workshops end not with a ranked list but with commitments. Future applications should build that step into the process itself, so that workshops end not with a ranked list but with commitments.

## List of abbreviations

CDC: Centers for Disease Control and Prevention;
COREQ: Consolidated Criteria for Reporting Qualitative Research;
EDN: Electronic Disease Notification system;
EMR: electronic medical record;
IFHP: Interim Federal Health Program;
IME: immigration medical examination;
IOM: International Organization for Migration;
IQR: interquartile range;
IRCC: Immigration, Refugees and Citizenship Canada;
MH2C: Migration Health and Humanitarian Collective;
NARHC: North American Refugee Health Conference;
NGT: nominal group technique;
REB: Research Ethics Board;
REPRISE: Reporting Guideline for Priority Setting of Health Research;
SD: standard deviation;
WHO: World Health Organization.

## Declarations

### Ethics approval and consent to participate

The study was approved by the University of Calgary Conjoint Health Research Ethics Board (REB 18-2074). Survey respondents read a waiver of consent at the start of the survey, and completion of the survey was taken as implied consent. Workshop participants were informed that workshop outputs would be analysed and reported and provided verbal assent to participate. All methods were carried out in accordance with the Declaration of Helsinki.

### Consent for publication

Not applicable. No identifying information about individual participants is reported; survey quotations are attributed by respondent’s role only.

### Availability of data and materials

The survey instrument, codebook, ranked solution lists and workshop breakout summaries are provided as Additional files 1–6. De-identified open-ended survey responses and ranking tallies are available from the corresponding author on reasonable request, subject to approval by the University of Calgary Conjoint Health Research Ethics Board.

### Authors’ contributions

GF, AC, ML and NES conceptualized and coordinated the priority-setting workshop, developed the workshop design in partnership with IRCC, and oversaw its implementation. EN and DA analyzed NGT data during workshop. ACS conceptualized the qualitative analysis and developed the codebook and lead the manuscript production. ACS, PK, MYE and EN analysed the data and drafted the initial manuscript contributing equally, PK conceptualized and designed figures, while all authors (GF, AC, ML, NES, ACS, EN, MYE and PK) made significant contributions to subsequent reviews and read and approved the final manuscript. GF was responsible for submitting the manuscript for publication.

## Data Availability

All data produced in the present study are available upon reasonable request to the authors. The survey instrument, codebook, ranked solution lists, workshop breakout summaries, and coded problem and solution quotes are provided in the supplementary material (Additional files 1 through 6). De-identified open-ended survey responses and ranking tallies are available from the corresponding author on reasonable request, subject to approval by the Conjoint Health Research Ethics Board of the University of Calgary.

## Acknowledgements

We thank the workshop participants for their time and expertise, and in particular the refugee community leaders who opened the workshop by sharing their experiences of displacement and resettlement. We thank Immigration, Refugees and Citizenship Canada for co-hosting the workshop and the North American Society of Refugee Healthcare Providers for allowing it to be held ahead of the 2023 North American Refugee Health Conference; and the O’Brien Institute for Public Health and the School of Public Policy at the University of Calgary for their support. We thank Lisa Peterman for facilitating the workshop in close collaboration in close collaboration with Myles Leslie.

## Use of artificial intelligence

Generative artificial intelligence was used at one stage. Before the workshop, ChatGPT (GPT-3.5; OpenAI) assisted the research team in grouping similar survey-generated solutions and merging duplicates to form the preliminary solution list presented at Step 3. All groupings were reviewed and finalized by the research team, and participants reviewed and expanded the list at the workshop, so the candidate solutions that went to ranking were generated by participants. AI was not used in qualitative coding or analysis. AI was not used for the writing of the manuscript except for proof-reading for spelling and grammar.

## Supplementary Files

**Additional file 1.**

File format:.pdf.

Title: Ranked solutions by Resettlement phase: 1a pre-migration (60 solutions), 1b post-arrival (43), 1c long-term (38); marks workshop-generated solutions.

Description: List of all proposed solutions and their rankings. This includes solutions gathered through the asynchronous online pre workshop survey and from during the workshop itself as part of the idea expansion and clarification stage – NGT Step 3. Data is available for ranked proposed solutions for each Resettlement Phase.

**Additional file 2.**

File format:.pdf.

Title: Pre-workshop survey instrument: sections, questions, response formats and variables.

Description: Online asynchronous survey handed out to participants prior to the in-person workshop (NGT Step 1-2). Response formats for each question and variables are also included.

**Additional file 3.**

File format:.pdf.

Title: Codebook: the three-dimensional coding matrix.

Description: Codebook used to complete the qualitative coding of the online asynchronous pre-workshop survey responses (responses provided to questions with free text answers). Codebook includes definitions of the three Resettlement Phases and the Stances (problems, barriers, enablers and solutions) used for coding.

**Additional file 4.**

File format:.pdf.

Title: Workshop breakout discussion by priority: 4a Resettlement phase 1, 4b Resettlement phase 2, 4c Resettlement phase 3.

Description: Summary of notes taken during the breakout group discussion during the workshop (NGT Step 5a). Groups discussed three questions across each station: 1. what is working well that can be leveraged?; 2. What is not working and can be improved?; and 3. How can we enhance coordination?. Data for all three Resettlement Phases is provided.

**Additional file 5.**

File format:.pdf.

Title: Problems and barriers by Resettlement phase: 5a pre-migration to initial post-arrival, 5b early post-arrival, 5c long-term resettlement. Themes, subthemes, and illustrative quotes.

Description: All quotes coded as “problems” or “barriers” by the coding team, from the online asynchronous pre-workshop survey and the in-person workshop discussions. Quotes are organized by inductive subthemes, created by coding team consensus.

**Additional file 6.**

File format:.pdf.

Title: Solutions proposed in the survey by Resettlement phase: 6a pre-migration to initial post-arrival, 6b initial post-arrival, 6c long-term resettlement. Themes, subthemes, and illustrative quotes.

Description: All quotes coded as “enablers” or “solutions” by the coding team, from the online asynchronous pre-workshop survey and the in-person workshop discussions. Quotes are organized by inductive subthemes, created by coding team consensus.

**Additional file 7.**

File format:.pdf.

Title: Reporting checklist (REPRISE).

Description: Completed REPRISE checklist

