## Additional File for "Priorities to Commitments: A binational adapted-NGT for refugee health prioritization"

**Additional file 1.** Ranked solutions by Resettlement phase: 1a pre-migration (60 solutions), 1b post-arrival (43), 1c long-term (38);  marks workshop-generated solutions.

**Additional file 1a. Ranked solutions, Resettlement phase 1 (pre-migration and early post-arrival): 60 candidate solutions**

| Solution | 1st (5) | 2nd (4) | 3rd (3) | 4th (2) | 5th (1) | Total points |
|---|---|---|---|---|---|---|
| 1. Connect pre-departure IME exams and health assessments to Canada’s refugee clinics. | 7 | 3 | – | – | 1 | 48 |
| 19. Allocate federal funding to improve equity in health care delivery. | 5 | 2 | 1 | 2 | 1 | 41 |
| 47. As refugee voices are yet to influence provincial and federal governments to move policy, engage with private sector champions of refugee health who will work with refugees themselves to move up agenda.* | 3 | – | 3 | 3 | 1 | 31 |
| 10. Create a task force of stakeholders for ongoing planning and coordination, involving Primary Care Networks, AHS, and Immigrant/Refugee serving agencies. | 2 | 5 | – | – | – | 30 |
| 13. Provide improved education for clinics and hospitals not specialized in newcomer health. | 1 | 2 | 3 | 3 | 1 | 29 |
| 52. Improve understanding and health care provider education of unique trauma and cultural/social contexts of refugee communities to create adaptable services.* | – | 3 | 3 | 3 | 2 | 29 |
| 15. Enhance communication and coordination between immigration authorities, healthcare providers, and refugees for timely sharing of medical information. | 2 | 3 | 1 | 1 | – | 27 |
| 33. Implement governance models to enforce co-planning and delivery across different levels and sectors. | – | 1 | 5 | 1 | 2 | 23 |

| | | | | | | |
|---|---|---|---|---|---|---|
| 2. Provide an online portal for accessing IME results. | 2 | 2 | 1 | – | – | 21 |
| 22. Strengthen communication between IRCC and agencies providing refugee health services. | 1 | 1 | 3 | – | 1 | 19 |
| 42. Standardize access to interpretation along journey from pre-migration, during migration and to post-migration. | 2 | – | 1 | 1 | 3 | 18 |
| 56. Engage in evidence-based, collective advocacy with government to treat root causes rather than symptoms. | 1 | 1 | 1 | 2 | – | 16 |
| 21. Implement inter-agency patient navigation and communication plans among agencies. | 2 | – | 1 | – | 1 | 14 |
| 45. Provision of coordinated healthcare responses for urgent issues when there is a rapid influx of refugees to cities, prior to refugee arrivals at hotels.* | – | – | 4 | 1 | – | 14 |
| 32. Increase guidelines and competency frameworks, conducting training in best practices. | 1 | 1 | 1 | – | 1 | 13 |
| 24. Broaden the scope of health assessments to include family systems and previous trauma. | 1 | 1 | 1 | – | – | 12 |
| 44. Develop methods for refugees to access information in country of origin in own language, e.g. video-based and easily accessible.* | 1 | 1 | – | 1 | 1 | 12 |
| 8. Increase funding and capacity for refugee screening clinics to conduct timely assessments. | – | 3 | – | – | – | 12 |
| 17. Enhance linkage of pre-migration information to resettlement agencies. | – | 3 | – | – | – | 12 |

| | | | | | | |
|---|---|---|---|---|---|---|
| 5. Improve communication between MDs conducting pre-medicals and healthcare teams in Canada. | 2 | – | – | – | – | 10 |
| 12. Foster strong baseline relationships between institutions through regular meetings for effective collaboration. | 1 | – | 1 | 1 | – | 10 |
| 53. Formal government mandates for settlement agencies to designated primary care sites, with funding enablers and resources attached for implementation support.* | 1 | – | – | 1 | 2 | 9 |
| 59. Look at barriers within Interim Federal Health Program.* | – | – | 1 | 2 | 2 | 9 |
| 60. Research best practices among resettlement agencies in navigating/coordinating client care.* | – | 1 | – | 2 | 1 | 9 |
| 58. Educate refugees and provide awareness about Interim Federal Health Program benefits.* | 1 | – | – | – | 3 | 8 |
| 34. Create partnership models involving key stakeholders for improved patient care. | – | 1 | – | 1 | 2 | 8 |
| 43. Develop adequate and appropriate pre-migration triage information of incoming medical conditions.* | – | 1 | 1 | – | 1 | 8 |
| 55. Beyond primary care, provide funding and interpretation support so it follows patients to specialty and complex care.* | – | 1 | – | 2 | – | 8 |
| 7. Automate document and data flow for easy access among relevant parties. | 1 | – | – | 1 | – | 7 |
| 20. Enhance cross-sector collaboration and information sharing between government departments. | 1 | – | – | – | 2 | 7 |
| 57. Determining roles of stakeholders in settlement process and ensuring all stakeholders | 1 | – | – | 1 | – | 7 |

| | | | | | | |
|---|---|---|---|---|---|---|
| at the table to share perspectives and improve coordination.* | | | | | | |
| 4. Ensure individuals receive their IME results to share with primary care providers. | – | 1 | 1 | – | – | 7 |
| 49. Consistent funding for refugee claimants.* | – | 1 | – | 1 | 1 | 7 |
| 25. Establish a medical screening program for newly arrived refugees and prepare newcomers about the healthcare system and processes. | – | 1 | – | 1 | – | 6 |
| 26. Designate care plans for complex patients. | – | – | – | 3 | – | 6 |
| 30. Work towards EMR interoperability and clear mapping of responsibilities. | – | – | 1 | 1 | 1 | 6 |
| 36. Communication and awareness of these issues to the public.* | – | – | 2 | – | – | 6 |
| 6. Establish a centralized database for investigations, blood test results, and physician assessments. | 1 | – | – | – | – | 5 |
| 11. Facilitate care coordination using community health workers, especially for insurance-reimbursable services. | 1 | – | – | – | – | 5 |
| 40. Re-envisioning how we use health assessment time: developing sub-criteria; using time to catch up on standard series vaccinations.* | 1 | – | – | – | – | 5 |
| 51. Leverage existing expertise to help with vaccine backlog.* | 1 | – | – | – | – | 5 |
| 9. Partner with local clinics to enhance access to timely evaluations. | – | 1 | – | – | 1 | 5 |
| 23. Conduct more research focused on the stressors experienced by refugees to improve assessment measures. | – | – | 1 | – | 2 | 5 |

| | | | | | | |
|---|---|---|---|---|---|---|
| 29. Use technology and apps to improve coordination of paperwork and services. | – | – | 1 | 1 | – | 5 |
| 31. Provide dedicated training and support for panel physicians. | – | – | 1 | 1 | – | 5 |
| 14. Ensure proper sharing of pre-departure field-collected health data with domestic teams. | – | 1 | – | – | – | 4 |
| 16. Improve communication and transfer of health records access. | – | 1 | – | – | – | 4 |
| 18. Support and standardize healthcare at international and federal levels. | – | 1 | – | – | – | 4 |
| 28. Encourage electronic transmission of health information for refugees and increased financial support for others. | – | – | 1 | – | 1 | 4 |
| 38. Standardized education and training resources for providers in host country clinics and university programs. | – | – | – | 2 | – | 4 |
| 41. Due to differing standards between countries of origin and host countries, be cognizant of access issues to migration to refugee resettlement countries.* | – | – | – | 1 | – | 2 |
| 48. Treating iCARE as a real-time profile of refugees' journey/as a proxy for understanding the refugee journey.* | – | – | – | 1 | – | 2 |
| 50. Data forecasting for future decision-making rather than retrospective data gathering only.* | – | – | – | 1 | – | 2 |
| 35. Improve understanding of service scope and promote better data sharing practices. | – | – | – | – | 1 | 1 |
| 39. Develop a portable health card or record for refugees to facilitate initial health assessments.* | – | – | – | – | 1 | 1 |

| | | | | | | |
|---|---|---|---|---|---|---|
| 46. Beyond triage, coordinate responses for refugees with physical disabilities to ensure their needs are met.* | – | – | – | – | 1 | 1 |
| 54. Advance settlement agencies'/workers' understanding of health issues, triage and cross-sector collaboration through new research.* | – | – | – | – | 1 | 1 |
| 3. Standardize expectations and capacity for optimizing pre-migration health assessments. | – | – | – | – | – | 0 |
| 27. Implement streamlined and easily accessible computer systems for domestic users. | – | – | – | – | – | 0 |
| 37. Standardized overseas history and examination training.* | – | – | – | – | – | 0 |

Values are the number of participants who placed each solution at each rank; weights in parentheses. Total points = sum of rank counts × weights (629 across all solutions). Solutions are listed by total points; numbering reflects the order in which they were generated.

*Generated at the workshop rather than drawn from the survey.

" –" = none.

**Additional file 1b. Ranked solutions, Resettlement phase 2 (early post-arrival, up to two years): 43 candidate solutions**

| Solution | 1st (5) | 2nd (4) | 3rd (3) | 4th (2) | 5th (1) | Total points |
|---|---|---|---|---|---|---|
| 14. Implement multidisciplinary care coordination, covering primary care, mental health, employment, legal, and housing services. | 3 | 4 | 1 | 4 | 1 | 43 |
| 11. Provide dedicated funding and support for health system navigation. | 3 | 1 | 4 | 2 | 1 | 36 |

| | | | | | | |
|---|---|---|---|---|---|---|
| 2. Establish cross-trained health navigators with expertise in settlement, healthcare, and mental health. | 4 | 2 | – | 1 | 3 | 33 |
| 37. Normalizing refugee health care as a component of primary healthcare and providing adequate supports for same* | 1 | 2 | 4 | 4 | – | 33 |
| 8. Expand the capacity of existing refugee clinics or establish more branches to support refugees. | 1 | 5 | 1 | 1 | – | 30 |
| 26. Better data flow between organizations* | 2 | 1 | 1 | 4 | 2 | 27 |
| 7. Create a mechanism to scale up health supports during refugee surges, including integrated health supports at hotels. | – | 4 | 2 | 1 | 3 | 27 |
| 3. Ensure sustained funding for healthcare initiatives. | 3 | 2 | – | 1 | – | 25 |
| 22. Chronic disease management* | 3 | 2 | – | – | 1 | 24 |
| 30. More direct contact between pre-departure and post-arrival* | – | 2 | 2 | 3 | 1 | 21 |
| 16. Ensure all Government Assisted Refugees (GARs) are connected to primary care services within 2-4 weeks after arrival. | 1 | 2 | 1 | 1 | 1 | 19 |
| 21. Trauma literacy among those providing care to refugees* | 1 | 1 | 2 | 1 | 2 | 19 |
| 1. Provide universally accessible translation services for mental health providers. | 3 | – | 1 | – | – | 18 |
| 18. Maintain ongoing interpretation services regardless of healthcare coverage. | – | 3 | 1 | 1 | 1 | 18 |
| 35. Alternate mental health models that are refugee-led* | – | 1 | 1 | 3 | 5 | 18 |
| 34. Training healthcare providers across all health specialties about working with refugees* | 2 | 1 | – | 1 | 1 | 17 |

| | | | | | | |
|---|---|---|---|---|---|---|
| 6. Offer extensive training in refugee health and mental health. | 1 | 2 | 1 | – | 1 | 17 |
| 25. Cross-sector collaboration to address competing priorities of refugees (school, work, appointments)* | 1 | 1 | 2 | 1 | – | 17 |
| 28. Leveraging IMGs to decrease healthcare burden and capacity issues† | – | – | 4 | 2 | 1 | 17 |
| 42. Create training for refugees to become cultural brokers (“training of trainers” model)† | 2 | – | – | 1 | 4 | 16 |
| 9. Foster communication and collaboration between community organizations, clinics, hospitals, and public health departments to identify needs and find solutions for new groups. | 2 | – | 1 | 1 | – | 15 |
| 43. Exploring various models of refugee welcome centres, and what that looks like for different kinds of refugees (e.g. GARs, claimants, etc.) to improve long-term job success* | 2 | – | – | 1 | 3 | 15 |
| 17. Offer incentives for primary care providers to take on refugee patients. | 1 | 1 | 1 | 1 | 1 | 15 |
| 29. Training on competency standards for healthcare workers* | 1 | – | 1 | 2 | 2 | 14 |
| 13. Increase availability of free and affordable mental health services for refugees of all ages. | 1 | – | 2 | – | 2 | 13 |
| 10. Form partnerships with community organizations to enhance support for refugees’ health needs. | 1 | 1 | 1 | – | – | 12 |
| 31. Automatic access to interpretation services* | – | – | 3 | – | 2 | 11 |
| 38. Ensuring that complex cases are settled to geographic areas with required support and resources* | – | 2 | 1 | – | – | 11 |

| | | | | | | |
|---|---|---|---|---|---|---|
| 4. Strategize and plan for future health services in a sustainable manner with partner involvement. | 1 | 1 | – | – | – | 9 |
| 40. Develop systems-level tools (e.g. provider standards, billing codes, accreditation) to enable providers to routinely use interpretation services* | – | – | 1 | 3 | – | 9 |
| 15. Engage clinicians in supporting resettlement agencies. | 1 | – | 1 | – | – | 8 |
| 39. Attaching funds to complex cases to incentivize providers to care for higher need patients* | 1 | – | – | 1 | 1 | 8 |
| 20. Incorporating screening and preventive care into refugee care* | – | 2 | – | – | – | 8 |
| 23. Social determinants of health when addressing refugee health and how it impacts our partnerships* | – | – | 2 | – | 2 | 8 |
| 19. Specific research and QI* | 1 | – | – | – | 2 | 7 |
| 33. Self-organized refugee community groups by and for refugees* | – | – | 1 | 2 | – | 7 |
| 24. Accessibility and appropriateness of IFH and ensuring it meets refugee needs. | – | 1 | – | 1 | – | 6 |
| 12. Involve settlement service providers and community partners in health initiatives. | 1 | – | – | – | – | 5 |
| 27. Better integration of different subsets of refugees (GARs, claimants, evacuees, etc.)* | – | 1 | – | – | – | 4 |
| 32. Leverage AI to tackle complex care needs/problems in refugee space* | – | – | 1 | – | 1 | 4 |
| 41. Fund refugee-led initiatives to help overcome social determinants of health barriers* | – | – | – | 1 | 1 | 3 |

| | | | | | | |
|---|---|---|---|---|---|---|
| 5. Implement a team-based setting for refugee health with adequate resourcing for interpretation. | – | – | – | 1 | – | 2 |
| 36. Incorporate gender-specific programming in the host country to facilitate transition* | – | – | – | – | – | 0 |

Values are the number of participants who placed each solution at each rank; weights in parentheses. Total points = sum of rank counts × weights (669 across all solutions). Solutions are listed by total points; numbering reflects the order in which they were generated.

*Generated at the workshop rather than drawn from the survey.

“–" = none.

**Additional file 1c. Ranked solutions, Resettlement phase 3 (long-term resettlement): 38 candidate solutions**

| Solution | 1st (5) | 2nd (4) | 3rd (3) | 4th (2) | 5th (1) | Total points |
|---|---|---|---|---|---|---|
| 4. Standardize national protocols and tools to manage influxes of refugees during humanitarian crises. | 9 | 1 | 1 | – | 1 | 53 |
| 28. Fully fund public health.* | 4 | 3 | 1 | 1 | 1 | 38 |
| 8. Increase funding for care coordinators and interpretation services in clinics serving refugees. | 4 | 3 | 1 | – | – | 35 |
| 32. Universal access to interpreter services guaranteed by institutional accreditation and national funding for it.* | 3 | 4 | 2 | 4 | 3 | 48 |
| 19. Build capacity in both primary care settings and refugee screening clinics. | 3 | 4 | 2 | 3 | 3 | 46 |
| 33. Readiness of refugee health clinics to respond to surges of new arrivals.* | 3 | 3 | 2 | 2 | 1 | 38 |

| | | | | | | |
|---|---|---|---|---|---|---|
| 3. Secure sustainable and less siloed funding. | 3 | 2 | – | – | 2 | 25 |
| 31. Address staff burnout by increasing capacity and retention.* | 2 | 1 | 2 | 1 | 6 | 28 |
| 23. Increase number of healthcare professionals and physicians.* | 2 | 1 | 3 | 1 | – | 25 |
| 21. Equal access to health care regardless of status: create a system for categories of arrivals who do not fit the UN convention definition of “refugee”.* | 2 | 2 | – | 3 | – | 24 |
| 35. Develop clearly established protocols for emergencies to facilitate implementation and de-implementation.* | 2 | 1 | 2 | – | – | 20 |
| 27. Single-payer healthcare system.* | 2 | 1 | – | – | 2 | 16 |
| 10. Invest in infrastructure, workforce development, and training to address staffing shortages. | 1 | 2 | 1 | 2 | 1 | 21 |
| 22. Strong data management and reporting.* | 1 | 1 | 1 | 3 | 2 | 20 |
| 24. Improve transition out of refugee health clinics into primary care.* | 1 | 1 | 1 | 3 | 1 | 19 |
| 5. Strengthen coordination between IRCC and provincial health systems. | 1 | 2 | 1 | 1 | – | 18 |
| 13. Establish stronger relationships within newcomer communities to understand and address their needs. | 1 | – | 2 | 2 | 2 | 17 |
| 37. Create a learning forum for ongoing dialogue and discussion about best practices.* | 1 | – | 2 | – | 4 | 15 |
| 26. Address pre-existing inequalities and disproportionate impact of Covid-19 on vulnerable communities.* | 1 | – | 1 | 2 | – | 12 |

| | | | | | | |
|---|---|---|---|---|---|---|
| 30. Look at how to scale up to meet acute needs in healthcare crises.* | 1 | – | 1 | 1 | – | 10 |
| 11. Foster collaboration among healthcare providers, government agencies, and community organizations. | – | 3 | 5 | – | 1 | 28 |
| 6. Ensure stable federal funding for refugee health clinics. | – | 3 | 2 | 1 | 3 | 23 |
| 17. Treat refugees, asylum seekers, and evacuees equitably in healthcare access. | – | 1 | 1 | 5 | 4 | 21 |
| 20. Develop communication plans with key partners to improve coordination. | – | – | 4 | 2 | 1 | 17 |
| 34. Embed clinical expertise within resettlement agencies to create bridge between overseas and host country.* | – | 1 | 1 | 3 | 3 | 16 |
| 2. Improve communication between different stakeholders. | – | 2 | 1 | 1 | – | 13 |
| 29. Advocacy around enabling policy for healthcare systems and refugee health.* | – | 1 | 1 | 1 | 1 | 10 |
| 25. Examine and improve timeline for approval in Interim Federal Health program.* | – | 1 | 1 | 1 | – | 9 |
| 14. Stay flexible and adaptable to handle larger volumes of newcomers with little notice. | – | 1 | – | 2 | – | 8 |
| 38. Encourage allied healthcare professionals to learn about the Interim Federal Health program and how to sign up for it.* | – | – | 1 | 1 | 3 | 8 |
| 15. Bring healthcare services to settlement agencies. | – | 1 | 1 | – | – | 7 |
| 12. Prioritize patient-centered care and timely access to services. | – | 1 | – | 1 | – | 6 |

| | | | | | | |
|---|---|---|---|---|---|---|
| 16. Develop specialized healthcare services for refugees. | – | – | – | 1 | 2 | 4 |
| 9. Provide oversight and monitoring to prevent clinics from being overwhelmed. | – | – | 1 | – | – | 3 |
| 18. Prioritize staff who are linguistically and culturally concordant. | – | – | 1 | – | – | 3 |
| 1. Implement team-based care. | – | – | – | – | – | 0 |
| 7. Enhance support for specialist services (e.g., optometry and audiology) billing under IFH. | – | – | – | – | – | 0 |
| 36. Different payment options for non-critical care based on socioeconomic status.* | – | – | – | – | – | 0 |

Values are the number of participants who placed each solution at each rank; weights in parentheses. Total points = sum of rank counts × weights (704 across all solutions). Solutions are listed by total points; numbering reflects the order in which they were generated.

*Generated at the workshop rather than drawn from the survey.

"–" = none.

**Additional file 2.** Pre-workshop survey instrument: sections, questions, response formats and variables.

**Pre-workshop survey instrument: sections, questions, response formats and variables**

**A. Respondent characteristics**

| # | Question (as asked) | Response format |
|---|---|---|
| 1 | What gender do you identify as? | Categorical |
| 2 | What was your sex at birth? | Categorical |
| 3 | How old are you? (age in years) | Numeric |
| 4 | You may belong to one or more ethnic or cultural groups on the following list. Which do you identify with? (select all that apply; "Other", specify) | Categorical, multiple |
| 5 | Name of your organization | Free text |
| 6 | Where is your organization located? (city; province/state; country) | Free text |
| 7 | What type of agency or organization is it? (if primary care: what best describes your primary care practice?; "Other", specify) | Categorical with free text for each option: federal government; provincial/state government; municipal/local government; primary care; specialty care; public health; settlement/resettlement agency; international organization; non-governmental organization (NGO); university or academic institution; other (please specify). If primary care: conditional item on the type of practice, with "other" free text |
| 8 | How frequently do you interact directly with refugees, asylum seekers or evacuees as part of your work? | Categorical: daily; weekly; monthly; rarely |

| | | |
|---|---|---|
| 9 | In what capacity do you usually interact with refugees, asylum seekers, and/or evacuees? (select all that apply; "Refugee Community Leader/Representative" and "Other", specify) | Categorical, multiple: healthcare provider (doctor, nurse, therapist, etc.); interpreter/cultural liaison; policy development or administration; refugee community leader/representative; administrator or researcher; settlement/resettlement agency; other |

**B. Resettlement phase 1: pre-migration and early post-arrival**

| # | Question (as asked) | Response format |
|---|---|---|
| 1 | How do you perceive access to pre-departure health assessment and medical history information for refugees, asylum seekers, and/or evacuees post-arrival for domestic healthcare providers? (0 = extremely difficult to access; 10 = extremely easy to access) | 0–10 scale (difficulty), with "other" option |
| | Please explain | Free text |
| 2 | What solutions can improve pre-migration and early post-arrival [screening]: a. that works well | Free text |
| | b. that needs improvement | Free text |
| 3 | How can successes in healthcare delivery and inter-agency integration and coordination [be amplified or scaled to other jurisdictions]? | Free text |
| 4 | What solutions can improve healthcare delivery and inter-agency integration and coordination? | Free text |

**C. Resettlement phase 2: early post-arrival, up to two years**

| # | Question (as asked) | Response format |
|---|---|---|
| 1 | To the best of your knowledge, how accessible are mental health services for newly arrived refugees, asylum seekers, and evacuees in your jurisdiction? | 0–10 scale |
| | Please explain | Free text |
| 2 | Please identify areas of post-arrival refugee healthcare services and integration with settlement:<br>a. that work well | Free text |
| | b. that need improvement | Free text |
| 3 | How can successes in addressing post-arrival refugee healthcare services and integration with settlement be amplified or scaled to other contexts? | Free text |
| 4 | What solutions can improve post-arrival refugee healthcare services and integration with settlement? | Free text |

**D. Resettlement phase 3: long-term resettlement**

| # | Question (as asked) | Response format |
|---|---|---|
| 1 | How do you perceive the capacity of existing health and settlement services in your jurisdiction in relation to current numbers of refugee, asylum seeker, and evacuee arrivals? (0 = extremely insufficient; 10 = extremely sufficient) | 0–10 scale |
| | If more than 5, please explain | Free text |
| 2 | How do you perceive the capacity of existing health and settlement services in your jurisdiction in relation to projected numbers of refugee, asylum seeker, and evacuee arrivals? (0 = extremely insufficient; 10 = extremely sufficient) | 0–10 scale |
| | If ≤5, please explain | Free text |

| | | |
|---|---|---|
| 3 | Please identify areas of healthcare and public health systems: a. that successfully adapt to large volumes of refugees, asylum seekers, and evacuees (i.e., resilient systems) | Free text |
| | b. that need improvement | Free text |
| 4 | How can successes in resilient healthcare and public health systems be amplified or scaled to other jurisdictions? | Free text |
| 5 | What solutions can improve areas of healthcare and public health systems resilience? | Free text |

**Additional file 3.** Codebook: the three-dimensional coding matrix.

**Resettlement phase**

| Resettlement phase | Working definition | Boundary test |
|---|---|---|
| Pre-migration and early post-arrival (≈ 0–6 months) | The migration journey and the handoff: preparing to move, moving, and making first contact with the receiving health system, including health assessments conducted before departure, the records they generate, whether that information travels with or ahead of the person, and the initial connection to care after arrival. | Does this concern the journey itself, or the handoff into first care? The phase ends where routine care in the community begins. |
| Early post-arrival (to ≈ 2 years) | The settlement window: ongoing care in the community while the person remains attached to specialized or transitional services, including referrals, coverage in practice, communication across language in the clinical encounter, and the eventual transition into the mainstream system. | Is the subject a person's ongoing receipt of care during the settlement window, while still attached to specialized or transitional services? |
| Long-term resettlement (2+ years) | The long run and the system as a system: health beyond the settlement window, after transition into mainstream care, and how the system is designed, funded, staffed and prepared so that it performs durably, including under surges of arrival. | Is the subject either health and care beyond the settlement window, or the system as architecture? A complaint about current rules blocking current care belongs to the settlement window, however national it sounds. |
| Overarching | Content belonging to the whole journey rather than any stretch of it: a statement whose claim runs across the phases as one continuous condition, experience or requirement. | Would filing this in a single phase misrepresent it? Overarching is not a fourth period; it marks continuity. |

**Stance**

| Stance | Working definition | Diagnostic question |
|---|---|---|
| Problem | A deficit, gap, undesirable condition, or challenge, stated without an accompanying fix or action. | What is wrong or missing? |
| Barrier | An operational mechanism, condition, or process that prevents or obstructs a solution, desired outcome, or access to care. | What is blocking improvement or preventing the solution from working? |
| Proposed solution | A proposed action, intervention, change, or improvement intended to address a problem or barrier. | What should be done? |
| Enabler | Something already happening that supports access, coordination, adaptation, or another positive outcome. | What is currently working well? |
| Context | Information about the setting, population, time, place, or circumstances without a clear judgment, obstacle, or proposed action. | Is this mainly background needed to understand the situation? |

**Additional file 4.** Workshop breakout discussion by priority: 4a Resettlement phase 1, 4b Resettlement phase 2, 4c Resettlement phase 3.

**Additional file 4a. Workshop breakout discussion by priority, Resettlement phase 1 (pre-migration and early post-arrival)**

| Priority | What is working and can be leveraged | What is not working and how to improve it | How to enhance integration and coordination |
|---|---|---|---|
| **Connect pre-departure IME exams and health assessments to Canada's refugee clinics** | EDN as a replicable model; IOM–EDN collaboration overseas | Arrivals lack their own records; tests are repeated or not performed; IMEs miss some conditions; Can be improved by providing a unique identifier for arrivals | Add individual health needs to IMEs for triage on arrival; Share overseas information with post-arrival assessments; Transition the interim health plan to provinces; Work with NGOs collecting data at border crossings; Enhance IFH; Technology-based solutions |
| **Allocate federal funding to improve equity in healthcare delivery** | Partnerships with Public Health and creation of provincially funded public health programs (e.g., routine vaccination) | Sustainable funding for continuous infrastructure development, training, and resource allocation. Funding should be specific for health and migration rather than compete for other programs. | Regular coordination meetings; Research on which models work, and funding for them |
| **Recruit private sector champions, in** | Influencers with lived experience to share | Improve public narrative on refugees; private sector and | Educate sectors on the value brought by refugees; |

| | | | |
|---|---|---|---|
| **collaboration with people with lived experience of refugee health*** | their stories on social media | refugee Community collaboration; Integration of internationally trained physicians | Internationally trained physicians as mentors for refugee care |
| **Create a taskforce of all key stakeholders for ongoing planning and coordination** | Relationship based Same-sector collaboration (e.g., health and shelter); Establishment of councils, convenors and partnerships for PSR's; Yearly refugee health policy conference; IRCC health liaison | Multi-sector collaboration is harder to achieve; Communication gaps pre- and post-migration; Need to improve role clarity, using an SDOH lens and improving language skills | Funded taskforce; Direct connection with IRCC; Refugees included in planning; Provider–agency partnerships; Centres of excellence (IRCC, PHAC, CHRN); Continue hosting yearly conference |
| **Provide improved education for clinics and hospitals not specialized in refugee health** | Specialized refugee clinics in newcomer hotspots; accessible refugee health resources; shared best practices | Provider training, especially on IFH; events and conferences to share knowledge | Training for administrators and leaders; interprofessional dialogue; better triage for primary care attachment |

Condensed from the research team's summaries of the breakout station notes (workshop report, Appendix C). Priorities are listed in rank order

* Solution generated at the workshop rather than drawn from the survey.

**Additional file 4b. Workshop breakout discussion by priority, Resettlement phase 2 (early post-arrival, up to two years)**

| Priority | What is working and can be leveraged | What is not working and how to improve it | How to enhance integration and coordination |
|---|---|---|---|

| | | | |
|---|---|---|---|
| **Implement multidisciplinary care coordination** | Expanding more training for providers with limited experience working with refugees. Improving mental health care through multidisciplinary care | Language barriers while providing care. Limited mental health support through the public mental health system. Limited funding for refugee health programs. | Partnerships between local hospitals and agencies with sustainable funding. Creation of specialized refugee medical homes such as the Burnaby clinics that provide different support for clients. |
| **Health navigation (dedicated funding and support for navigation; cross-trained health navigators)** | Care models that include behavioral paths within their care, such as the Minnesota clinic; Health navigators, counseling groups, and partnerships between providers and agencies. | Lack of health navigators and family doctors. Difficulty navigating the health system. Lack of rule clarity between providers and agencies. | Training health navigators to play their crucial roles. Providing culturally appropriate services, and role clarity between settlement agencies and providers |
| **Normalize refugee health care as a component of primary health care*** | Using the community to map clinics with access to medical interpreters and referring refugees to associated family physicians. | Lack of support for Ukrainians navigating the Canadian health system. Making resources for practitioners and specialists more user-friendly and more easily accessible. | Use websites and apps such as the Refugee Care App (Canada) Care Ref (US) to provide resources for general practitioners, specialists and providers to access information, especially during screenings. |
| **Expand the capacity of existing refugee clinics** | Increasing the number of specialists and translation services. Access to mental health services supported by | Limited availability of counseling and interpretations services. Family physicians not accepting new | Increasing partnerships between settlement agencies and providers for more outreach and educational services. |

| | | | |
|---|---|---|---|
| | community organizations such as the CCIS and Center for Refugee Resilience. | patients, lack of providing culturally appropriate services and limited funding. | Recruiting more healthcare providers trained in refugee health. Establishing a network of providers who can speak other languages as well. Sustainable fundings. |
| **Better data and flow of information between organizations*** | Ensuring electronic medical records exist for each refugee and standardization of the records. | Lack of data sharing agreements affecting the data flow between the IRCC and providers. Limited data flow between settlement agencies and the health system. Difficulty tracking client data beyond Year 1 of arrival, after they leave the refugee clinic. | Developing models of data flow. Using community-based health navigators to engage the community when we need to collect data and disseminate information. |

**Additional file 4c. Workshop breakout discussion by priority, Resettlement phase 3 (long-term resettlement)**

| **Priority** | **What is working and can be leveraged** | **What is not working and how to improve it** | **How to enhance integration and coordination** |
|---|---|---|---|
| **Readiness for surges: national protocols and tools to manage influxes; readiness of refugee health clinics** | Standardize national protocols to allow health and settlement agencies to scale up resources. | Inadequate staff due to lack of funding. Difficulty meeting the needs of arrivals due to increasing pace of arrivals. Not enough primary care doctors with sufficient training | Expand healthcare infrastructure to address capacity limitations. Enhance regulatory oversight to prevent demand exceeding capacity. More specialized refugee health services. |

| | | | |
|---|---|---|---|
| | | and experience in refugee health | |
| **Guarantee universal access to interpretation services*** | Recurring more staff who are linguistically and culturally concordant; Engage local public health systems in screening and immunization. Engage settlement agencies to help refugees navigate post-arrival health screenings; Billing IFH to hire more providers and see more patients, and extending IFH beyond 1 year | Appointments cancellation due to lack of interpretation services, not enough physicians speaking other languages or lack interpretation support. High interpretation cost; More educational outreach for refugees to understand healthcare navigation, comprehensive coverage, two separate payment systems; Delays and no increase in funding to meet the budget and staff, limited coordination between provincial and federal | Streamline billing and increase the number of providers. More efficiency in primary care for refugees such as language support and decreasing administrative barriers; Embedding health coordinators directly within both resettlement agencies and medical clinics, valuing lived experience within care coordination roles, increasing funding and incentive for refugee health professionals. |
| **Build capacity in both primary care settings and refugee screening clinics** | Passion and interest in dedicated refugee clinic good;work gets done bad because of burnout/unpaid work. Formalize interprofessional care Public clinics, serving under served (US model) refugee and non-refugee care | Formalize inter professional care Learning from each other Inclusion/Exclusion criteria Coordinate care of vulnerable/under served | Operationalize what works well throughdata/research. Education in all health care professionals in refugee care. Define Efficiency in primary care for refugee care. Resources for all primary care languages. Decrease administration to simplify. Define exactly what is needed so we can easily and |

| | | | |
|---|---|---|---|
| | | | succinctly tell funders. Define integration and funding. |
| **Fully fund public health / single payer system*** | “Comprehensive” and easy access for patients Fairly straightforward for clinicians Strong public support Strong brand Cost-effective | Navigation of services Education for refugees Certain services (e. PT, OT), excluded More comprehensive coverage Funding for additional public health programs to improve access Two payment systems (IFH and provincial) Coordinators to help bridge gap Importance of policy for advocacy | Extend access to care by additional training, payment, etc. to individuals beyond MDs, etc. Integrate systems ex. IFH and provincial system to streamline billing and increase providers |
| **Increase funding for care coordinators in clinics serving refugees** | Some health care systems pay for care coordination (ORR). Funding for health care through universal health coverage. Health related case management by settlement agencies. | Lack of funding for care coordination and a limited ability to bill for it. Gaps in care coordination for privately sponsored refugees and other claimants. | Integrating health coordinators within both resettlement agencies and medical clinics. Integrating inter-agency communication pathways. |

**Additional file 5.** Problems and barriers by Resettlement phase: 5a pre-migration and early post-arrival, 5b early post-arrival, 5c long-term resettlement. Themes, subthemes, and illustrative quotes.

**Additional file 5a. Problems and barriers, Resettlement phase 1 (pre-migration and early post-arrival): themes, subthemes, and illustrative quotes**

| Subtheme | Illustrative quote |
|---|---|
| Information blocked or delivered inconsistently between settlement agencies and health clinics | *"It appears that whatever information is available is accessible for the settlement agency, but does not necessarily get transferred to refugee health clinics."* (Primary care) |
| | *"I do not receive pre-departure health assessments or histories. Information I receive is only anecdotal."* (Provincial/state government) |
| | *"Unless someone makes an effort to bring their documentation with them, it is unlikely to become part of their new health record - this feels like a real luxury or highly unusual situation!"* (University/Academic institution) |
| | *"On a local level, we have worked closely on our relationship with the local settlement agency and still the information received is inconsistent."* (Primary care) |
| | *"Asylees and some humanitarian parolees lack documentation and/or it's not included in an easy-to-access system. The infrastructure to house the data isn't there."* (Provincial/state government) |
| Data is not captured or updated systematically or consistently, resulting in incomplete medical history | *"Lack of updated medical information not available on EDN."* (Primary care) |

| | |
|---|---|
| | *"Early post arrivals - We do not receive IME but we receive Notification Arrival Transmission (NATs) sometimes will indicate health needs (eg dialysis, person care, physical/mental health needs)."* (Settlement /Resettlement agency) |
| | *"Rap providers receive basically one sentence of information when there is something serious - that notification is only to ensure the client get timely access to service."* (Settlement /Resettlement agency) |
| | *"Inconsistent, sometimes we have access to NATS or patients bring pre-departure medicals, more often we don't have access to anything even for patients with significant health conditions i.e. 90% there are no record, 10% there is nothing."* (Primary care) |
| Data is not centralized; providers cannot access information consistently and rely on patients bringing their own records | *"It is very rare for healthcare workers to access any pre-departure medical exam or tests."* (Primary care) |
| | *"Extreme duplication in the system is created due to lack of pre-arrival information, health assessment included."* (Primary care) |
| | *"Sometimes information is not there (eg dialysis) We know more about the clients (blood pressure, diabetes) when clients talk to us or we ask needs assessment and clients disclose their health issues so we can access services needed."* (Primary care) |
| | *"it is very difficult for healthcare providers to access this information and depends on clients having access to and bringing their own health records (which may or may not be in English)."* (Primary care) |

| | |
|---|---|
| Medical records not sent or received with significant delays, resulting in clinical delays, long referral times or inability to plan | *"Sometimes we are blindsided with the needs of the client early post-arrival."* (Settlement /Resettlement agency) |
| | *"Also the NATS are general or received a few days before arrivals so unable to prepare."* (Settlement /Resettlement agency) |
| No formal coordination or navigation channels between federal immigration agencies and state/provincial health clinics | *"For regular arrivals, there are no regular channels for communication between IRCC and health care providers."* (Primary care) |
| | *"The current system seems very fragmented with multiple stakeholders that do not have readily available channels to communicate with each other. This does impact healthcare delivery within the early resettlement. Undoubtedly."* (Primary care) |
| | *"We cannot see what the other agencies are doing with patients real-time, and this hampers coordination and progress."* (Municipal/local government) |
| No formal information transfer pathways between federal immigration agencies and state/provincial health clinics | *"There is not a process for IRCC to forward information to health care providers - only some to federal public health who then inform provincial public health when there is a serious concern."* (Settlement /Resettlement agency) |
| | *"Given the transfer of pre-departure medical information between those administering it and federal/provincial services is not guaranteed, it is apt to get lost."* (Academic/ University Institution) |
| | *"Patient may come with medical export and then nothing advised after arrival to Canada."* (Primary care) |

| | |
|---|---|
| Specialized migrant and refugee clinics are scarce; services are over-saturated, resulting in prolonged waiting times and delayed care | *"In the cases where there are no designated primary care providers, the refugee may have to wait long before an assessment is done to really assess their health needs."* (Settlement /Resettlement agency) |
| | *"Many parts of Canada do not have refugee health clinic or designated primary care services for refugees and rely on walk in clinics and ER initially."* (Settlement /Resettlement agency) |
| | *"But it takes time to get a provincial healthcare number which can delay some tests/treatment."* (Primary care) |
| | *"Wait times can be long and hard to get clients connected to medical services/supports."* (Settlement agency) |
| | *"Mental health - a very long referral process and so a very long time before clients see a mental health professional."* (Settlement /Resettlement agency) |
| Health professionals with limited training in context and specific migrant-refugee needs may be limited in their ability to identify and address health needs | *"Identification of children with complex needs."* (Primary care) |
| | *"Vagueness of what services clients need and we are not equipped/trained/suitable to provide high level of medical needs. We do not have 24/7 medical supports."* (Settlement /Resettlement agency) |
| | *"Unrealistic expectations on the NAT - ie 2 wk appt for neurologist, 1 wk for cardiologist."* (Primary care) |

| | |
|---|---|
| Low supply of clinicians and clinics that accept federal limited health coverage versus high demand | *"Not doctors accept IFH and those who do accept, the medical offices do not know how to process IHF."* (Settlement/Resettlement agency) |
| Lack of specialized or sensitive health services available | *"Lack of medical interpretations including not having languages that our clients need and create delays. Our staff cannot provide medical interpretation."* (Settlement /Resettlement agency) |
| Budgetary constraints impacting infrastructure development, recruitment, retention and training of health personnel | *"There is still a struggle to get enough staffing resources both at the clinic and in the RAP SPOs to respond to the increase in GARs. the funding always lag. We see staff burnout and large turnover - these issues reduce access to timely services for clients."* (Settlement /Resettlement agency) |
| | *"Most refugee clinics operate on minimal budgets and can not hire extra case management staff."* (Primary care) |
| | *"Resettlement agency support - have noted infrastructure still lacking since funding had been cut under Trump administration, needs are higher than response capacity for resettlement agencies particularly for larger unplanned migration of groups (ex. Afghans)."* (Academic/ University Institution) |

**Additional file 5b. Problems and barriers, Resettlement phase 2 (early post-arrival, up to two years): themes, subthemes, and illustrative quotes**

| Subtheme | Illustrative quote |
|---|---|
| Coordination between sectors remains a challenge; lack of delimitation of roles and responsibilities | *"Actors in the space are often underfunded, and do not have the bandwidth (for coordination), personnel (to coordinate) or means to move staff to meeting sites."* (Municipal/Local Government) |

| | |
|---|---|
| | *"It has been over 10 years since the model of health care was developed. Need to regroup to strategize and plan for health services in the future. Our current model is not sustainable and does not involve multiple partners."* (Provincial/ State Government) |
| | *"Minimal communication with the mental health service providers and primary care providers."* (Primary Care) |
| | *"Needs improvement – Care coordination for those with significant medical conditions and mental health issues."* (Federal Government) |
| Lack of linkage between IRCC, settlement agencies and healthcare providers | *"For regular arrivals, there are no regular channels for communication between IRCC and health care providers."* (Primary Care) |
| | *"No connection between pre arrival clinic where I did all necessary tests and Canadian immigration system."* (N/A) |
| Specialized services limited by the low and declining number of available and eligible clinicians; health navigation challenges | *"Health system navigation support is needed and challenges to attachment to a regular family MD."* (Primary Care) |
| | *"For migrants/refugees in administrative detention other services are available, but not to the standard available in the community."* (Municipal/ Local Government) |
| | *"Casual racism is a concern as well (e.g. many providers were more willing to offer support for Ukranians and less so for Afghans)."* (Provincial/ State Government) |
| Mental health services are scarce and uneven | *"There is very limited access to mental health services this is compounded by language equity issues and expertise in refugee or immigrant experience."* (Primary Care) |

| | |
|---|---|
| | *"Services are variable. Some clinics have excellent access while for others it is almost impossible to obtain psychiatric care or quality counselling."* (Primary Care) |
| | *"Most services are only available if there is hospitalization - sometimes counselling or group sessions can support clients enough to help with them with settling."* (Settlement/ Resettlement Agency) |
| | *"Refugees often require specific mental health supports that are not readily available through the public mental health system."* (Primary Care) |
| | *"Services are behind gatekeepers (e.g. physicians), but are accessible; so long as folks are not in detention."* (Municipal/ Local Government) |
| | *"Almost inaccessible for asylum seekers but a little easier if GARS and PSR's evaluated within our network of clinics."* (Primary Care) |
| Within scarcity, culturally sensitive mental health services that respond to needs are unavailable | *"Services may be concentrated in particular parts of the state or providers may have limited serving refugees/immigrants."* (Provincial/ State Government) |
| | *"Limited affordable or free counselling available in general let alone in first language and/or cultural sensitivity."* (Settlement/ Resettlement Agency) |
| | *"Our state is working on this but our mental health capacity and resources are limited, especially when considering culturally appropriate services."* (Provincial/ State Government) |

| | |
|---|---|
| | *"Mental health workers in the region of little background in trauma and violence informed care with a lens of the refugee experience."* (Provincial/ State ) |
| Cultural and linguistic needs may complicate quality | *"Sometimes cancel appointments if not interpretation available."* (Settlement/ Resettlement Agency) |
| | *"Language barriers alone make accessing mental health services, almost impossible in the general medical system."* (University/ Academic Institution) |
| | *"Clients who do not speak English and able to access medical interpretation are not served."* (Settlement/ Resettlement Agency) |
| Funding landscape challenges scaling up successful models | *"We used to have Transcultural Mental Health Program at our local hospital but this folded several years ago due to lack of funds."* (Primary Care) |
| | *"Very challenging due to limited services at baseline for all state, county, city residents and then added shortage of providers that are linguistically and culturally concordant."* (University/ Academic Institution) |
| Underfunding leaves staff doing work outside their role, including finding doctors who cover | *"Not all doctors accept IFH so workers spend time looking for doctors that accept IFH and/or accept new patients - taking away the role of providing settlement supports."* (Settlement/ Resettlement Agency) |
| | *"Difficulty affording over the counter medications on arrival."* (Primary Care) |
| Unmet health needs displace settlement priorities | *"Clients are not focused on settlement if they are dealing with health issues or have health needs."* (Settlement/ Resettlement Agency) |

| | |
|---|---|
| | *"I saw lots of Ukrainians coming and not receiving necessary support from health care services and settlement organizations. This is one of the biggest reasons why lots of them prefer to go back to Ukraine."* (NA) |

**Additional file 5c. Problems and barriers, Resettlement phase 3 (long-term resettlement): themes, subthemes, and illustrative quotes**

| Subtheme | Illustrative quote |
|---|---|
| Large influx of individuals with complex needs exacerbates chronic challenges in meeting demand for local and refugee populations | *"Our waitlist for our clinic has been up to 200 for a prolonged period of time (recently received funding and have increased our capacity)."* (Primary Care) |
| | *"Many systems are unable to accommodate large numbers of arrivals and see them in a timely manner, this is further worsened by lack of capacity in volags and permanent housing availability."* (Primary Care) |
| | *"If current healthcare system has issues with wait times, lack of family physicians for all those already in Canada and not able to increase medical services, how will it be possible to support more clients with the existing services?"* (Settlement/ Resettlement Agency) |
| | *"Resettlement agencies are having difficulties meeting the needs of newcomers for various reasons: increased pace of arrivals, complex medical needs, inability to increase their capacity (case managers' salaries are very low and it's a stressful job)."* (Provincial/ State Government) |
| | *"I believe, Canadian system is too overloaded with all refugees that are coming and lots of services need to solve existing* |

| | |
|---|---|
| | *problems in the order to provide necessary care for future refugees."* (Primary Care) |
| Limited human and financial resource capacity to meet needs, compounded by staff burnout and turnover | *"Not enough primary care doctors, acute care system not able to handle the needs of all projected refugees, lack of providers with the appropriate skills and training to provide support to refugees."* (Primary Care) |
| | *"Not enough family physicians accepting new patients, enough doctors who speak first language or willing to access provincial medical interpretation or accept IFH patients."* (Settlement/ Resettlement Agency) |
| | *"(Severe shortage of resources and personnel) Driven by mismatch between federal funding and the influx of immigrants/refugees."* (NA) |
| | *"We are currently only funded for 1NP and 1RN for refugee health from the province. All other resources are 'borrowed' from the community health centre. This limits our ability to grow and we are forced to turn down supporting many others who are in need of health supports (primary care and allied health)."* (Provincial/ State Government) |
| | *"We have 70 000 people in our city without family physicians. We don't have any primary care providers to pass patients on to after they have been seen and had immediate needs met at our refugee health clinic."* (Primary Care) |
| | *"Professionals within the health and settlement sector are experiencing burnout as a result of vicarious trauma, the covid-19 pandemic, access to appropriate resources* |

| | |
|---|---|
| | *to meet client/patient needs, the health human resource crisis, etc."* (Other) |
| | *"We are unable to get projected numbers so unable to plan."* (Primary Care) |
| | *"We also struggle with access to interpretation funds. Even at our CHC interpretation costs are over $250 000 per year."* (Primary Care) |
| Funding has not kept pace with demand, affecting operations, staffing and retention | *"We are using provincial money to provide care for the refugees that the federal government decides to bring to our city."* (Primary Care) |
| | *"Financial support for clinics that support these groups has not increased to match."* (Public Health) |
| | *"Funding has not increased to meet demand, and retention among settlement organizations is low as a result."* (Other) |
| | *"But without adequate funding and resources, these approaches are not sustainable in the long-term."* (Other) |
| | *"Resources are available, but the model is not efficient or coordinated."* (Provincial/State Government) |
| | *"IFH is still a service that still have challenges. we are still seeing delays and refusal when there are no other coverage and in very complex cases. there should be no reason for having to do heavy handed advocacy to get things covered... IFH is still being run like a regular medical insurance where they are trying to save money - the focus should be more about providing* |

| | |
|---|---|
| | *coverage based on the need of the client."* (Settlement/ Resettlement Agency) |
| | *"It is not easy for specialists and independent primary care providers to bill IFH for interpretation so sometimes referrals are refused."* (Primary Care) |

**Additional file 6.** Solutions proposed in the survey by Resettlement phase: 6a pre-migration and early post-arrival, 6b early post-arrival, 6c long-term resettlement. Themes, subthemes, and illustrative quotes.

**Additional file 6a. Solutions proposed in the survey, Resettlement phase 1 (pre-migration and early post-arrival): themes, subthemes, and illustrative quotes**

| Subtheme | Illustrative quote |
|---|---|
| Centralize and/or transfer blood test results, imaging and clinical assessments | *"Should receive actual imaging results such as CT chest to prevent repeating testing upon arrival."* (Public Health) |
| | *"Centralized database for all investigations, blood test results, and physician assessments where we can look our patients up - i.e. TB screening CXR, HIV test. NAATs could even be included on this database."* (Primary Care) |
| | *"Should receive actual documents/diagnostic images, not an interpreted narrative of those results."* (Public Health) |
| Leverage existing platforms to develop a safe and continuous process for medical records to be streamlined within and across jurisdictions | *"If there was a centralized resource for medical record distribution, or better channels for this communication this preparation could happen more seamlessly flowing from pre-departure or IOM medical screening to primary care resources in the community, and then referral onto specialized care where needed."* (Primary Care) |
| | *"Better integration of the iCare reporting system with the Interim Federal Health Plan would allow better linkage and communication between settlement agencies working on the cultural, economic, and social inclusion of refugees and those providing direct care. This would permit a 'whole-of-person' approach to their care, particularly in the first year."* (University/Academic Institution) |
| | *"Centralized portal to include IME that can be accessed by health care providers."* (Other) |
| Standardize what and how information is captured and shared within and across jurisdictions | *"An understanding and list of criteria for officers completing the NAT. For example, mobility troubles need to be included, medications; reason for urgent hospital attention etc. In all these situations, detail is required, not just a nice to have, however the NATs are often poorly filled and not descriptive enough to attend to patient needs."* (Municipal/Local Government) |
| | *"Need to send refugees to Canada with paper copies of pre-departure health assessments that are standardized in English and French and past medical history information that they can provide to domestic providers post arrival in Canada. This would bypass* |

| Subtheme | Illustrative quote |
|---|---|
| | *any privacy concerns as the patient themselves can consent to providing their health information to their new healthcare providers."* (University/Academic Institution) |
| Strengthen communication between agencies and sectors, and develop dissemination opportunities to share best practices | *"Need better coordination between agencies. Meetings are sporadic and reactive. Minimal opportunity for ongoing planning and coordination."* (Provincial/State Government) |
| | *"Through regular meetings or conferences, we can learn about best practices in other jurisdictions. IRCC can play a leadership role in ensuring equity of services across jurisdictions."* (Primary Care) |
| | *"Communication plans among agencies with key point people designated."* (Primary Care) |
| Engagement with local communities to build bottom-up solutions | *"Providing more outreach to refugees to explain health care systems in post-migration settings."* (Public Health) |
| | *"Ensure that medical cases have extensive local/community support (ie. have family or community ties)."* (Provincial/State Government) |
| | *"Creating platforms that recognizes refugees as equals partners."* (University/Academic Institution) |
| Rather than operate in silos and by sectors, build strong cooperation within and across actors delivering health and social support for refugees | *"All agencies engaged in refugee resettlement need to be in communication, leveraging their respective resources (minimize duplication of efforts by coordinating services between agencies)."* (Provincial/State Government) |
| | *"We need a Canada wide approach that include key stakeholders (from a lens of social determinants of health) - there need to be a political will and leadership from the federal government and its departments and from the provincial departments."* (Settlement/Resettlement Agency) |
| | *"We need to create different partnership models to involve the key stakeholders to get the best care for our patients."* (Primary Care) |
| | *"Governance models that enforce co-planning and delivery across the divides and jealousies between: federal-provincial; provincial primary care and provincial public health; provincial-municipal; ngo/private contractors for settlement services and* |

| Subtheme | Illustrative quote |
| --- | --- |
| | *community base as well as health sector services."* (University/Academic Institution) |
| | *"Strengthening partnerships with stakeholders in public health as a mechanism for cross-sharing of emerging promising practices."* (Public Health) |
| Strengthen workforce capacity through training and dedicated linking roles | *"Dedicated training and support for panel physicians."* (Primary Care) |
| | *"Every settlement agency has a health co-ordinator to link to the refugee health clinic."* (Primary Care) |
| | *"By offering opportunities for training and education to refugee health care providers and professionals."* (Federal Government) |
| | *"Standardizing expectations and capacity to optimize the pre-migration health assessment would be helpful."* (Provincial/State Government) |
| | *"Sharing of relationship building techniques and 'how to' lessons from local initiatives that are succeeding with particular attention to policies (governance and resourcing)."* (University/Academic Institution) |
| Cross-sector awareness of roles and realistic expectations | *"More awareness of the resettlement process in medical centers that care for many refugees."* (University/Academic Institution) |
| | *"Physicians doing the Medicals need accurate expectations of the health care system and long wait times for specialist appointments."* (Primary Care) |
| | *"Understanding of our role as RAP providers and needing supports from healthcare on high needs, complex medical issues. We cannot support them and need more involved and understand the consequences if medical supports do not intervene earlier than when crisis happens. Often intervention happens during and after a crisis."* (Settlement/Resettlement Agency) |
| Knowledge on health system and how to access care | *"Clear information about Canadian healthcare in the official website. Non profit organizations who would explain about Canadian healthcare."* (Person with Lived Experience) |
| | *"Development of a website to disseminate these guidance materials along with brief (1-3 minutes) videos highlighting key success stories of how these challenges were overcome."* (Public Health) |

**Additional file 6b. Solutions proposed in the survey, Resettlement phase 2 (early post-arrival, up to two years): themes, subthemes, and illustrative quotes**

| Subtheme | Illustrative quote |
|---|---|
| Expand health coverage, either through expansion of the network or seed funding for specialized care (e.g., mental health) | *"Consider how [we] can provide funding to bolster local mental health supports."* (Primary Care) |
| | *"Primary care after 12 months (within the general population) as solo physicians do not have the time and or the supports (e.g. interpretation) to provide care."* (Provincial/ State Government) |
| | *"If we could bill IFH for each refugee we see this would provide us with our own basket of money protected for refugee health, we could hire more providers and see more patients."* (Primary Care) |
| | *"IFH needs to be extended beyond the 1 year for these types of patients because specialists wait lists are so long."* (Primary Care) |
| Ensure culturally and linguistically sensitive health services | *"Culturally concordant mental health care, weekly community based groups for social support in transition."* (University/ Academic Instution) |
| | *"Universally accessible translation services for mental health providers."* (University/ Academic Institution) |
| | *"Establishing a network of mental health clinicians that speak a languages other than English and/or French and have diverse cultural and racial identities."* (Other) |
| Expand refugee-sensitive health delivery (training, specialized services, health navigators) | *"Expanding training to providers who have limited experience working with refugees, especially in rural/mid-size cities where refugees/evacuees are settling."* (Provintial/State Government) |
| | *"Establishing more branches of refugee clinics or increasing their capacity to support refugees for at least one year, similar to my experience, would be beneficial."* (Other) |
| | *"Cross trained health navigators that are trained in both settlement, healthcare and mental health, not just settlement and cross cultural competencies, which is the norm today."* (University/Academic Institution) |
| Develop community-based interventions, including individualized guidance, navigation and alternative approaches to in-patient care | *"Outreach/education, and connection to affordable mental health services."* (Public Health) |
| | *"Alternative approaches to mental health care that involve the community may be appropriate for* |

| Subtheme | Illustrative quote |
|---|---|
| | *populations where mental illness is highly stigmatized."* (Other) |
| | *"Access to expert consultants who can provide individualized guidance on how to set up."* (Primary Care) |
| Strengthen partnerships across clinics, settlement and public health to bridge service provision and increase access | *"Partnerships with medical providers and agencies to get them better access to care."* (Primary Care) |
| | *"Involve settlement service providers and community partners especially since health isn't only physical/mental health but also social determinants of overall being."* (Settlement/Resettlement Agency) |
| | *"Encouraging bridges to be built between community based organizations serving newcomers, clinics and hospitals, as well as public health departments to be in communication particularly for new groups to quickly identify needs and determine solutions."* (University/ Academic Institution) |
| Broaden the model of care beyond specialized clinics | *"Need to streamline services and develop a model of care that includes many more partners in order to manage capacity with the increasing numbers of refugees arriving."* (Provincial/State Government) |
| | *"Mainstreaming more interprofessional service availability into the health system at large, when refugees have nowhere else to go."* (University/ Academic Institution) |
| Clarify roles and responsibilities between health and settlement staff | *"Consider making their responsibilities clearer or hiring/funding social workers who are trained in counsellor and responsible to their college."* (Primary Care) |
| | *"Need to delineate who provides counselling, social supports and psychiatry – and scope of organizations providing counselling vs. settlement."* (Primary Care) |

**Additional file 6c. Solutions proposed in the survey, Resettlement phase 3 (long-term resettlement): themes, subthemes, and illustrative quotes**

| Subtheme | Illustrative quote |
|---|---|
| Scaling up already successful models of delivery | *"A mechanism to scale up health supports during refugee surges."* (Primary Care) |

| Subtheme | Illustrative quote |
| --- | --- |
| | *"Would be good to expand it (collaboration with regional authorities) more beyond that period so more accessible."* (Settlement/ Resettlement Agency) |
| | *"Both dedicated refugee health clinics that inherently offer the interprofessional services typically needed, and public health systems that are designed and committed to care for underserved populations and their social determinants of health."* (University/ Academic Institution) |
| | *"Looking at models of care that can be replicated in other cities."* (Primary Care) |
| Establish day-to-day communication channels and long-term 'best-practice' forums for continuous improvement | *"Opportunities to share lessons learned and best practices in places where resettlement healthcare and public health are strong through case studies to reach practitioners who see newcomers commonly and also uncommonly to ensure needs are being met and gaps are closed."* (University/ Academic Institution) |
| | *"Consider establishing a platform for knowledge exchange and collaboration between jurisdictions to facilitate the sharing of best practices and lessons learned… By promoting cross-jurisdictional learning, other regions can adapt and implement successful strategies to enhance their own healthcare and public health systems."* (N/A) |
| Standardize the approach with minimum competencies, delivering services according to international guidance and recommendations | *"Continue to create and share standards, efficiencies, etc so that when capacity shifts we are not recreating the wheel. Public health is affected by politics and other responses, so whatever we can do to build upon the great work we have in place now, when funding exists and systems have been developed, will support us again."* (Provincial/ State Government) |
| | *"We need standardize national protocols and tools that allow health and settlement to scale up and down together, with appropriate resources and personnel to manage acute large, volume influxes of refugees in response to humanitarian crisis, which are predicted to continue and worsen due to climate change."* (University/ Academic Institution) |
| Anchor interventions at community level by leveraging local voices in leadership | *"Improved relationships within newcomer communities for a voice and leadership to come directly from the group to assist healthcare and public health to address challenges and create culturally responsive solutions that are grounded in their reality."* (University/ Academic Institution) |

| Subtheme | Illustrative quote |
|---|---|
| Coordination and oversight as key enablers for scaling effective solutions | *"Enhanced oversight by the healthcare section of the government: Ensure regulatory oversight and monitoring of clinics to prevent situations where patient demand exceeds capacity… Invest in workforce development: Prioritize training and recruitment to address staffing shortages and ensure a sufficient number of healthcare professionals to meet patient needs."* (N/A) |
| | *"Strengthen coordination: Foster collaboration among healthcare providers, government agencies, and community organizations to improve communication and streamline processes, ensuring efficient and effective delivery of healthcare services."* (N/A) |
| | *"Stable sources of federal funding for refugee health clinics. Distribution of complex cases that matches available resources. Communication directly with refugee health clinics. Also adding a channel for feedback for the clinics back to IRCC about what we are seeing on the ground… Consideration for funding for care co-ordinators and interpretation services for clinics that provide services to refugees."* (Primary Care) |
| | *"There needs to be increased coordination between IRCC and provincial health systems."* (Primary Care) |

Quotes in Additional files 5 and 6 are reproduced verbatim from survey responses, with minor punctuation added for readability; respondents are identified by role only.

### Additional file 7. REPRISE reporting checklist

Reporting guideline for priority setting of health research (REPRISE).

*Tong A, Synnot A, Crowe S, et al. BMC Med Res Methodol. 2019;19:243.*

#### Context and scope

| No | Item | How addressed in this study |
|---|---|---|
| 1 | Define geographical scope | Binational: Canada and the United States, with participants from six Canadian provinces, seven US states and international organizations. Priorities address federal, provincial and state levels and local service delivery. |
| 2 | Define health area, field, focus | Refugee and migrant health: healthcare delivery and health system organization across three resettlement phases (pre-migration to early arrival; post-migration to two years; long-term resettlement). |
| 3 | Define end-users of research | Forcibly displaced people in resettlement, defined to include refugees, asylum claimants and Ukrainian evacuees, and the health and settlement services that serve them. |
| 4 | Define the target audience of the priorities | Federal, provincial and state policymakers, health system and settlement sector leaders, clinicians and funders in Canada and the United States. |
| 5 | Identify the broad research area | Health services and health policy. This exercise set policy and service-delivery priorities rather than research questions, a departure from most REPRISE applications that is stated in the |

| No | Item | How addressed in this study |
|---|---|---|
| | | Methods. |
| 6 | Identify the type of research question | Not applicable. Priorities were proposed solutions for service delivery, coordination and policy, closest to health services and implementation. |
| 7 | Define the time frame | No interim, short-term or long-term labels were applied to the priorities. The separate actionability vote distinguished what existing actors could begin from what requires structural change. No schedule for revising the priorities was set. |

**Governance and team**

| No | Item | How addressed in this study |
|---|---|---|
| 8 | Describe selection of the leadership and management team | Planning group convened by the University of Calgary's Migration Humanitarian Health Collective (formerly Refugee Health YYC), the O'Brien Institute for Public Health and the School of Public Policy, in partnership with IRCC and the Society of Refugee Healthcare Providers. GF, AC, ML and NES conceptualized and coordinated the exercise and developed the design with IRCC. |
| 9 | Describe the characteristics of the team, and the networks they represent | Academic clinicians, health services researchers, a policy researcher and a project manager, based at the University of Calgary and working through the Migration Humanitarian Health Collective, with IRCC as federal co-host and the refugee healthcare provider society as conference host. |
| 10 | Describe any training or experience in priority setting | The workshop was led by an experienced external facilitator with expertise in organizational design and consensus processes. The research team had previously run an NGT priority-setting exercise with a refugee patient advisory |

| No | Item | How addressed in this study |
|---|---|---|
| | | committee. |

**Framework**

| No | Item | How addressed in this study |
|---|---|---|
| 11 | State the framework used (if any) | An adapted nominal group technique embedded within an appreciative inquiry framework, grounded in pragmatist and participatory epistemological stances. No established priority-setting framework such as the James Lind Alliance, COHRED or CHNRI was used. |

**Stakeholders and participants**

| No | Item | How addressed in this study |
|---|---|---|
| 12 | Define the inclusion criteria for stakeholders involved in priority-setting | Refugee community leaders, clinicians, academics, public health officials, policymakers and resettlement service providers from government, civil society and international organizations, invited to the workshop, the survey or both. |
| 13 | State the strategy or method for identifying and engaging stakeholders | Purposive and network sampling through established professional and community networks and registrants for the 2023 North American Refugee Health Conference. Invitations were sent by email between April and June 2023. |
| 14 | Indicate the number of participants and/or organisations involved | Sixty-eight people attended the workshop: 44 both days, 15 Day 1 only and 9 Day 2 only. Fifty-three completed the survey: 45 workshop participants and eight invitees who did not attend. |
| 15 | Describe the characteristics of stakeholders | Age, gender, country, sector and frequency of contact with refugees are reported in Table 1 for survey respondents and, through linked responses, for workshop participants. The 23 participants (34%) who did not complete the |

| No | Item | How addressed in this study |
|---|---|---|
| | | survey are counted as not reported. Numbers by stakeholder group are not reported. |
| 16 | State if reimbursement for participation was provided | **Confirmed:** no reimbursement was provided for participation. |

**Identification and collection of priorities**

| No | Item | How addressed in this study |
|---|---|---|
| 17 | Describe methods for collecting priorities from stakeholders | Asynchronous online survey as silent idea generation (Steps 1 and 2), with rating items and open-ended questions per phase on problems and barriers, what works well, how successes could be scaled and what solutions could improve care. At the two-day face-to-face workshop, participants reviewed the preliminary list and generated further solutions silently and individually (Step 3), added to the list in real time. |
| 18 | Describe methods for collating and categorizing priorities | The research team grouped similar survey solutions and merged duplicates into one preliminary list per resettlement phase. Generative AI (ChatGPT, GPT-3.5) assisted with that grouping; all groupings were reviewed and finalized by the team, and participants reviewed and expanded the list at the workshop. |
| 19 | Describe methods and reasons for removing priorities | Duplicate survey solutions were merged before the workshop. After the weighted ranking, the facilitator proposed combining solutions that described the same action and participants agreed in plenary which to merge; a merged priority took the rank of its highest-scoring component and each component's points are reported. Only the five highest-scoring priorities per phase went forward. Where two tied at fifth, participants chose as a group; in Phase 2 one tied solution was not carried to the final vote. |

| No | Item | How addressed in this study |
|---|---|---|
| 20 | Describe methods for refining or translating priorities into research topics or questions | Not applicable. Priorities remained policy and service-delivery solutions in participants' own wording, clarified through questions at Step 3 before ranking. |
| 21 | Describe methods for checking whether research questions or topics have been answered | Not done. No evidence review was conducted to establish whether the proposed solutions had already been evaluated elsewhere. |
| 22 | Describe number of research questions or topics | 141 candidate solutions, 73 from the survey and 68 generated at the workshop, reduced by weighted ranking to 15 leading priorities, five per phase. All 141 are listed with their points in Additional files 1a to 1c. |

**Prioritization**

| No | Item | How addressed in this study |
|---|---|---|
| 23 | Describe methods and criteria for prioritising research topics or questions | Step 4: weighted ranking of each participant's top five solutions per phase by confidential paper ballot, first to fifth choice receiving five to one points, with no criterion specified. Step 5a: progressive consensus building at stations structured by three appreciative inquiry questions. Step 5b: dot voting in plenary, one vote for the most important and one for the most actionable priority in each phase, in separate colours; a participant could choose the same priority for both. |
| 24 | Provide reasons for excluding research topics/questions | Priorities were carried forward on total weighted points, five per phase, a practical limit set by the two-day format rather than a threshold score; several lower-ranked solutions received substantial points. Because some ballots ranked fewer than five solutions and a few repeated a rank, the number of ballots per phase was taken as the largest number of votes |

| No | Item | How addressed in this study |
|---|---|---|
| | | cast at any single rank. |

**Output**

| No | Item | How addressed in this study |
|---|---|---|
| 25 | Specificity of research priorities are clear | Each of the 15 priorities is stated as a specific action, attributed to a resettlement phase, and reported with its weighted ranking points and its shares of most-important and most-actionable votes. |

**Evaluation and feedback**

| No | Item | How addressed in this study |
|---|---|---|
| 26 | Describe how the process of prioritization was evaluated | Station summaries were presented in plenary after each phase, followed by debate and member checking, and the 15 leading priorities were presented for comment in a closing plenary. |
| 27 | Describe the approach for feeding back priorities to stakeholders and/or to the public; and how feedback was addressed and integrated | Priorities were presented back to participants in the closing plenary, reported to IRCC as co-host, and set out in an internal workshop report produced in October 2023 and circulated to participants. |

**Implementation**

| No | Item | How addressed in this study |
|---|---|---|
| 28 | Outline the strategy or action plans for implementing priorities | No implementation strategy or action plan was established as part of the exercise. The Discussion sets out the implementation pathways the priorities imply, and the Conclusion argues that future applications should end with explicit commitments naming who will act, by when and with what resources. |
| 29 | Describe evaluation of impact | Not undertaken. The priorities were not followed after the workshop and implementation was not assessed. |

**Funding and conflict of interest**

| No | Item | How addressed in this study |
|---|---|---|
| 30 | State sources of funding | The workshop was supported by Immigration, Refugees and Citizenship Canada (IRCC), which also co-hosted it. Funds were administered through the Alberta Association of Immigrant Serving Agencies (AAISA) under project 10042228. |
| 31 | Outline the budget and/or cost | 65,490.00 CAD minus processing fees was assigned as grant for operations |
| 32 | Provide declaration of conflict of interest | Declared: the authors have no competing interests. |

NGT: nominal group technique. IRCC: Immigration, Refugees and Citizenship Canada. AAISA: Alberta Association of Immigrant Serving Agencies. NASRHP: North American Society of Refugee Healthcare Providers.
